# Developing an Early Diagnostic Signature and Deciphering the Microbial-Host Dynamics in Lower Respiratory Tract Infection (LRTI) in Paediatric Intensive Care Unit (PICU) Patients

**DOI:** 10.1101/2025.10.31.25335414

**Authors:** Zongtai Wu, Gehad Youssef, Iain R. L. Kean, Zhenguang Zhang, John A. Clark, Nazima Pathan, Namshik Han

## Abstract

**Background:** Lower respiratory tract infection (LRTI) is a leading cause of morbidity and mortality among children admitted to paediatric intensive care units (PICUs). Diagnosis is hampered by overlapping symptoms and limited sensitivity of conventional microbiology. We aimed to identify early diagnostic biomarkers by integrating microbial and host responses in paediatric LRTI.

**Methods:** We re-analysed a metagenomic next-generation sequencing (mNGS) dataset from 261 PICU patients with acute respiratory failure, combining microbial and host transcriptomic profiles using differential expression, network, and machine-learning approaches. Candidate biomarkers were validated in an independent prospective cohort of 100 critically ill children (RASCALS), with non-bronchoscopic bronchoalveolar lavage (mini-BAL), blood cytokine profiling, and pathogen detection.

**Findings:** Respiratory syncytial virus (RSV) and *Haemophilus influenzae* were the most enriched pathogens in LRTI cases. Host transcriptomics revealed activation of cytokine and chemokine signalling pathways. A seven-gene panel (IRF7, FFAR3, GZMB, FABP4, FN1, CXCL5, BCAR1) achieved high diagnostic accuracy, comparable to a published 14-gene model. In the RASCALS cohort, mini-BAL IL-1β, IL-4, and IL-8 classified bacterial LRTI with 65% accuracy, and blood IL-6 and TRAIL achieved 82% accuracy.

**Interpretation:** Integrating host and microbial markers provides a feasible route to early, accurate diagnosis of paediatric LRTI. The identified 7-gene panel and cytokine markers could be translated into PCR- or ELISA-based bedside assays to support rapid clinical decision-making and antimicrobial stewardship in PICU.

**Research in context:** *Evidence before this study:* LRTIs are a leading cause of morbidity and mortality in critically ill children^1^, yet conventional diagnostics often cannot distinguish true infection from colonisation, driving broad-spectrum antimicrobial use. Previous studies have generally examined host or microbial factors in isolation, leaving host–microbe interactions in ventilated patients poorly understood. *Mick et al.*^10^ showed that a 14-gene host signature could separate bacterial from viral infections, but did not address host–microbe dynamics. This highlighted the need for integrated diagnostic models in PICU.

*Added value of this study:* This study advances paediatric LRTI diagnostics by extending the analysis of the microbial and host inflammatory response using a machine learning and network analysis approach. Re-analysing 261 critically ill children from *Mick et al.*^10^ we identified respiratory syncytial virus (RSV) and *Haemophilus influenzae* as dominant pathogens and delineated immune pathways associated with disease progression. From this, we derived a seven-gene host biomarker panel (IRF7, FFAR3, GZMB, FABP4, FN1, CXCL5, BCAR1) that matched the performance of the 14-gene model but with greater simplicity. Network analyses revealed inflammatory pathways linked to RSV co-infections and prolonged ventilation. Importantly, we validated these findings in the independent RASCALS cohort, where IL-1β, IL-4, IL-8, IL-6 and TRAIL were associated with bacterial LRTI diagnosis and clinical outcomes (ventilator-free days).

*Implications of all the available evidence:* Our results support moving from pathogen-only diagnostics to integrative host–microbe models, particularly in mechanically ventilated children. RSV and *H. influenzae* emerge as major drivers of paediatric LRTI. The seven-gene host panel, together with cytokine markers identified through our network analysis, could be developed into rapid PCR- or ELISA-based point-of-care assays to guide antimicrobial decisions. Multi-omic diagnostics may allow earlier, more precise LRTI diagnosis and support antimicrobial stewardship. Further studies should test performance across diverse patient populations.

## 1. Introduction

Lower respiratory tract infection (LRTI) is a significant cause of morbidity and mortality among children globally, and the leading cause of death from infection in children under 5^1^. Severe LRTI is among the most common reasons for paediatric intensive care unit (PICU) admission, often requiring mechanical ventilation.^2^ Accurate diagnosis is difficult because of diverse aetiologies and overlapping presentations with non-infectious conditions.^3^ Conventional microbiology has limited sensitivity, as antimicrobial therapy is often initiated before samples are collected,^4^ leading to diagnostic uncertainty, empirical use of broad-spectrum antibiotics, and rising antimicrobial resistance.^5^

Standard microbiology is further constrained by slow culture times and the inability to detect atypical pathogens.^6^ Molecular approaches such as quantitative PCR and metagenomic next-generation sequencing (mNGS) broaden pathogen detection,^7^ while host transcriptomic profiling provides complementary insights into immune responses ^8,9^. Integrating host and microbial signals could enable earlier, more precise diagnosis of paediatric LRTI.

However, key gaps remain in understanding how specific pathogens, particularly viruses, interact with the host immune system to drive inflammation and tissue injury. Evidence increasingly suggests that host responses, rather than pathogen features, are the main drivers of LRTI pathophysiology. Further study of host–microbe interactions in ventilated patients is therefore needed.

To address this gap, we aimed to integrate microbial and host transcriptomic data to identify early diagnostic biomarkers and characterise disease mechanisms in paediatric LRTI. We analysed a published dataset from *Mick et al.*^10^, which includes metagenomic next-generation sequencing (mNGS) of pathogens, airway microbiome profiling, and host gene expression data from 261 children with acute respiratory failure in PICU.^11^ We then analysed data from an independent cohort of children with suspected LRTI (RASCALS study)^12^ to explore the pathogens and biomarkers identified. Our objectives were to: (i) identify microbial and host signatures as potential early diagnostic markers of LRTI; (ii) delineate host immune pathways associated with disease progression; and (iii) explore interactions between microbial colonisation and host responses in LRTI development.

## 2. Methods

### 2.1 Study Design and Participants

We obtained publicly available transcriptomic and clinical data from *Mick et al.*^10^, which included a prospective cohort of 261 mechanically ventilated children (31 days–17 years) admitted to eight PICUs within the National Institute of Child Health and Human Development’s Collaborative Paediatric Critical Care Research Network (2015–2017). The dataset comprised tracheal aspirate RNA-sequencing and associated clinical metadata from patients meeting the inclusion criteria of requiring invasive mechanical ventilation via an endotracheal tube for ≥72 hours. Further details on patient recruitment, ethical approvals, and sample collection procedures are available in *Mick et al.*^10^.

For the validation cohort, we obtained data from the Rapid Assay for Sick Children with Acute Lung Infection Study (RASCALS)^12^, a prospective cohort of 100 mechanically ventilated children under 18 years of age with suspected LRTI at Addenbrooke’s Hospital PICU in Cambridge, UK. Samples were obtained via non-bronchoscopic bronchoalveolar lavage (mini-BAL) and peripheral blood within 24 hours of enrolment for pathogen detection and for cytokine profiling. Final LRTI status was assigned retrospectively based on the primary diagnosis at hospital discharge, with diagnoses of bronchiolitis, pneumonia, pneumonitis, or other lower respiratory tract infections classified as LRTI, and all other diagnoses considered non-LRTI. Further details on patient recruitment, ethical approvals, and sample collection procedures are available in *Clark et al.*^12^.

### 2.2 LRTI Adjudication

LRTI diagnosis was adjudicated independently of mNGS results using (1) retrospective clinician review of all available clinical data and (2) standard-of-care respiratory microbiology from the first 48 hours of intubation.^8,11^ Patients were categorised as Definite (clinical and microbiologic evidence), Suspected (clinical diagnosis without microbiologic findings), Indeterminate (microbiologic findings without a diagnosis), or No Evidence (alternative non-infectious cause). For this study, we defined *Definite* cases as LRTI and *No Evidence* cases as controls.

### 2.3 Differential Expression and Microbial Abundance Analysis

Differential gene expression and differential microbial abundance were assessed using *DESeq2*^13^ (version 1.46.0) and *edgeR*^14^ (version 4.4.2), with normalisation via the median-of-ratios and TMM methods, respectively. Low-expression genes were filtered by excluding genes with fewer than 10 counts in less than 20% of samples prior to differential expression analysis. Genes were defined as differentially expressed based on a Benjamini-Hochberg adjusted P < 0.05 and absolute log₂ fold change > 2. Only genes consistently identified as significant by both *DESeq2* and *edgeR* were retained as final differentially expressed genes. Microbial abundance was analysed using *DESeq2*’s “poscounts” and edgeR’s TMMwsp methods, with significance at P < 0.1.

### 2.4 Protein Network Construction and Analysis

Differentially expressed genes were mapped to proteins using Ensembl Biomart.^15^ Protein-protein interaction networks were constructed using the STRING database^16^ with a confidence threshold of >0.4. Proteins not directly encoded by differentially expressed genes but connecting DEGs within the protein-protein interaction network (‘hidden layer proteins’) were included to capture intermediary proteins mediating host responses. Network centrality measures (eigenvector, degree, and betweenness centrality) and random walk with restart (RWR) were applied, with significance assessed via 1,000 permutations (P < 0.01).

### 2.5 Feature Selection and Machine Learning

Key proteins were mapped to gene IDs and analysed via LASSO logistic regression. Variance-stabilising transformation was applied using *DESeq2*^13^. Binary LRTI classifiers (Logistic Regression, Random Forest, SVM, k-NN, XGBoost) were trained with 5-fold cross-validation. Hyperparameters were optimised for ROC performance.

### 2.6 Pathway Analysis

Signaling pathway impact analysis^17^ (version 2.58.0) was conducted using the KEGG pathways provided within the package, incorporating over-representation and pathway perturbation metrics. Logistic regression classifiers were developed for each pathway, ranked by cross-validated F1 scores.

### 2.7 Meta-Analysis of RSV Studies

GEO datasets^18^ were screened up to August 2024 using the PICO framework^19^: children with respiratory infections (*P*), RSV exposure (*I*), healthy controls (*C*), and microarray-derived gene expression (*O*). Differential expression was analysed via GEO2R. Differentially expressed genes consistently identified in at least two of the four RSV studies were selected for integration into the meta-analysis.

### 2.8 Cytokine Classifier Development (RASCALS Validation)

In the RASCALS validation cohort^12^, we measured 48 cytokines in mini-BAL and plasma using the Bio-Plex Pro Human Cytokine Screening 48-plex kit. Clinical data were adjudicated by three PICU consultants to determine the presence of LRTI and classify infections as bacterial, viral, mixed or non-LRTI. For analysis, bacterial LRTI included both bacterial and mixed infections, non-bacterial LRTI referred to viral-only cases, and non-LRTI included patients without evidence of infection.

Logistic regression classifiers were trained separately using inverse-rank normalised mini-BAL and blood cytokine concentrations to distinguish bacterial LRTI from non-bacterial patients. Cut-offs were determined using the ‘closest to top-left’ criterion from receiver operating characteristic (ROC) curves. RSV status was incorporated into combined models for final performance evaluation.

### 2.9 Statistical Analysis

Differential expression and network significance were assessed as described above. Welch’s ANOVA, Welch’s t-test, Tukey’s test, and Kendall’s correlation were applied where relevant (P < 0.05 for tests, P < 0.1 for correlations).

### 2.10 Data and Code Availability

All data, code, and software packages used, including versions and citations, are available at https://github.com/zongtaiwu/mphil-paediatric-LRTI. The raw data, including patient adjudication, host gene counts, and microbial taxon counts were retrieved from *Mick et al.*^10^ at https://github.com/eranmick/pediatric-mNGS-LRTI-classifier. The code for protein network analysis was adapted based on the code from *Han et al.*^20^ at https://github.com/wchwang/COVID19.

### 2.11 Ethical and Regulatory Approvals

This study includes secondary analysis of clinical samples and data collected under the *Rapid Assay for Sick Children with Acute Lung Infection Study (RASCALS)*. The original RASCALS study received ethical approval from the Yorkshire and the Humber, Bradford Leeds Research Ethics Committee (Research Ethics Committee reference *20/YH/0089*), under the governance of the UK Health Research Authority (HRA). Ethical approval was granted for prospective participant recruitment, sample collection, and the use of routine and research-based diagnostic assays. Deferred consent for up to 48 hours was permitted in accordance with the approved protocol. For the current analysis, all samples and associated clinical data were anonymised prior to inclusion, and therefore no additional ethical approval was required. Informed consent was obtained from parents or legal guardians of all participating children enrolled in the RASCALS study.

## 3. Results

### 3.1 Dataset and Patient Selection

We first analysed microbial and host gene expression data from 261 children with acute respiratory failure admitted to PICU. Patients were categorised as Definite LRTI (n=117), No Evidence of LRTI (n=50), Suspected (n=57), or Intermediate (n=37). For primary analyses, Definite cases and No Evidence controls were included, while Suspected and Intermediate groups were excluded because of diagnostic uncertainty.

For validation, we examined a prospective cohort of 100 critically ill children with suspected LRTI (RASCALS). Each patient underwent non-bronchoscopic bronchoalveolar lavage (mini-BAL) and blood sampling for pathogen detection and cytokine profiling. Final diagnosis of LRTI or non-LRTI was assigned at hospital discharge and adjudicated by consultant consensus including classification as bacterial or non-bacterial infection. Clinical outcomes were recorded, with ventilator-free days at 28 days post-admission used as a marker of severity.recorded.

Figure SF1 summarises patient selection and study design.

### 3.2 Key Host Genes, Proteins, and Pathways Associated with LRTI Progression

After filtering low-expression genes, 13,323 transcripts were retained for host transcriptomic analysis. Unsupervised clustering and principal component analysis (PCA) showed clear separation between LRTI cases and controls, confirming distinct global expression profiles (Figures 1A–B).

**Figure 1.**
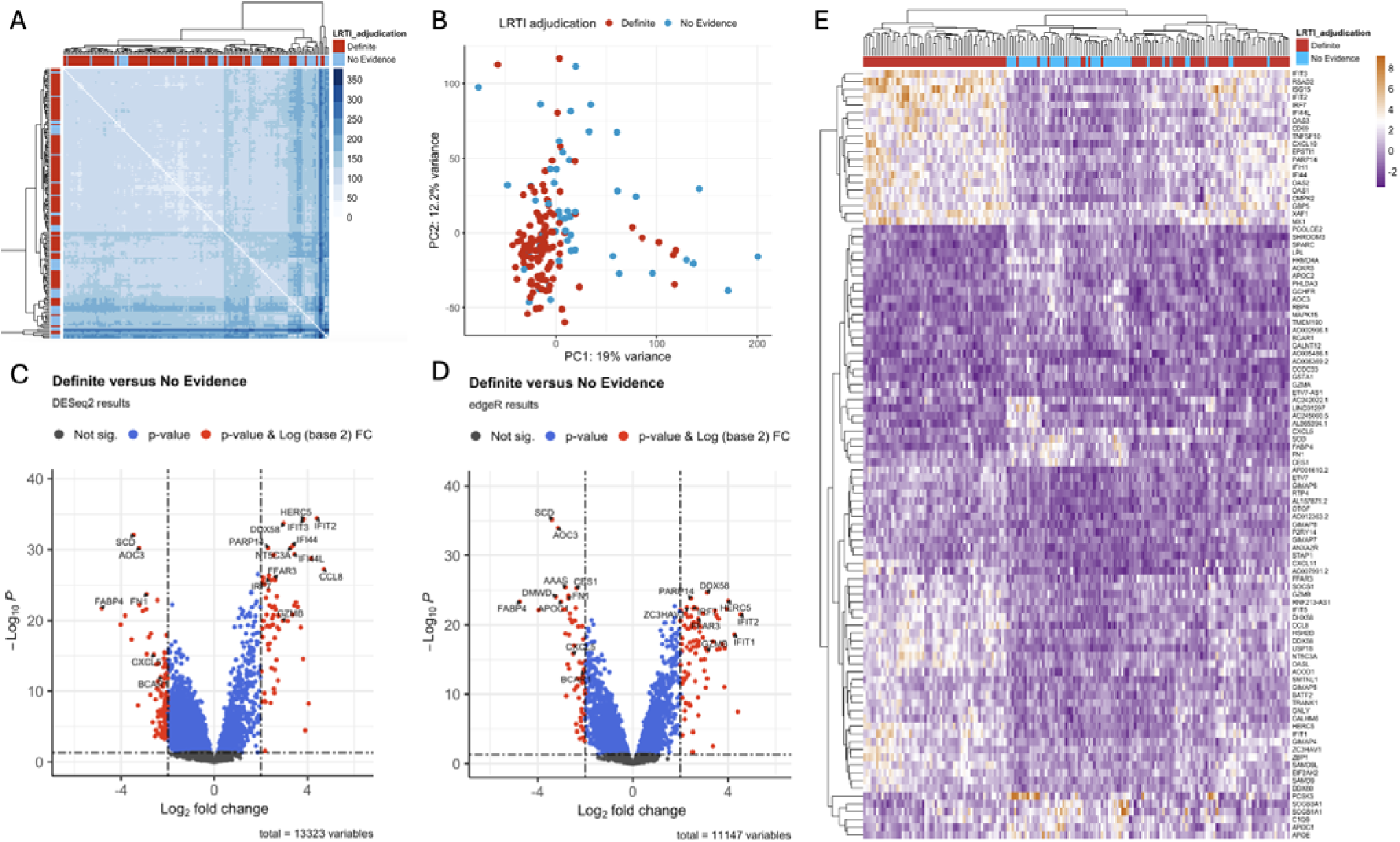
A) Heatmap of sample-to-sample Euclidean distances in LRTI samples and controls using the variance stabilising transformed (VST) values. B) PCA plot in in LRTI samples and controls using the VST values. C) Volcano plot of differential expression analysis between LRTI and control cohorts using DESeq2. FDR adjusted p-value < 0.05, and absolute log2 fold change > 2 was deemed significant. D) Volcano plot using edgeR. E) Heatmap of VST values across all patients from Definite and No Evidence (columns) for the 99 differential expressed genes intersecting from DESeq2 and edgeR (rows). LRTI adjudication (top colored horizontal bar) is displayed alongside the regression coefficients for each selected gene (side bar plot).

Differential expression analysis identified 180 genes by DESeq2 and 144 by edgeR, with 99 consistently detected by both methods (Figures 1C–D). These genes were enriched for antiviral and inflammatory responses, and hierarchical clustering showed robust separation between cases and controls (Figure 1E).

To assess functional relationships, we constructed a protein–protein interaction (PPI) network (Figure 2A). Network analysis highlighted 88 centrally positioned proteins (Figure 2B), suggesting key roles in host immune responses during LRTI.

**Figure 2.**
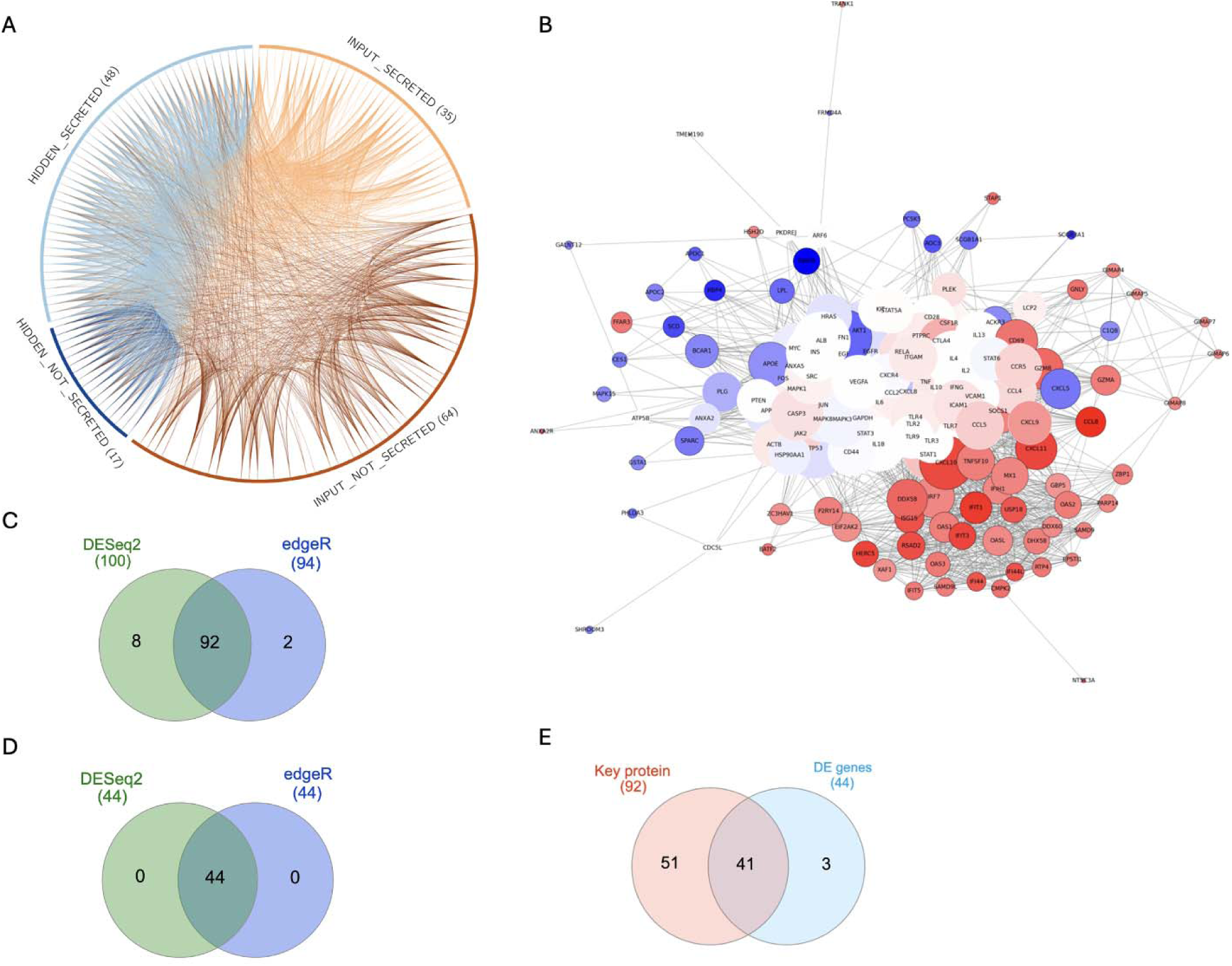
A) A circus plot depicting interactions between DEPs through the PPI network. The plot includes only input and key proteins, where 65 key proteins are in the hidden layer. The proteins are subdivided based on their secretion into bloodstream. The colored lines show PPI. B) A plot showing the network which includes only input and key proteins. Nodes with larger sizes have higher eigenvector centrality. The node colour reflects the log2 fold change in gene expression, with red indicating positive values, blue indicating negative values, and white indicating near-zero or N/A values. C) Number of pathways from SPIA using DESeq2 and edgeR results on key protein genes. D) Number of pathways from SPIA using DESeq2 and edgeR results on DEGs. E) Intersection of pathways in C) and D).

Pathway enrichment analysis revealed 41 consistently perturbed immune cascades, including cytokine, chemokine, Toll-like receptor, and interferon signalling (Figures 2C–E; Supplementary file).

Overall, LRTI was characterised by upregulation of inflammatory and antiviral pathways and activation of central network proteins likely driving disease progression.

### 3.3 LRTI Diagnostic Markers and Disease Pathways selected by Supervised Learning Methods

From 88 key network proteins, we derived a seven-gene diagnostic panel that distinguished LRTI cases from controls. The genes IRF7, FFAR3, and GZMB were upregulated, while FABP4, FN1, CXCL5, and BCAR1 were downregulated (Figures 3A–B). Two genes (CXCL5 and FABP4) overlapped with the previously reported 14-gene classifier,^10^, while five were unique to this study.

**Figure 3.**
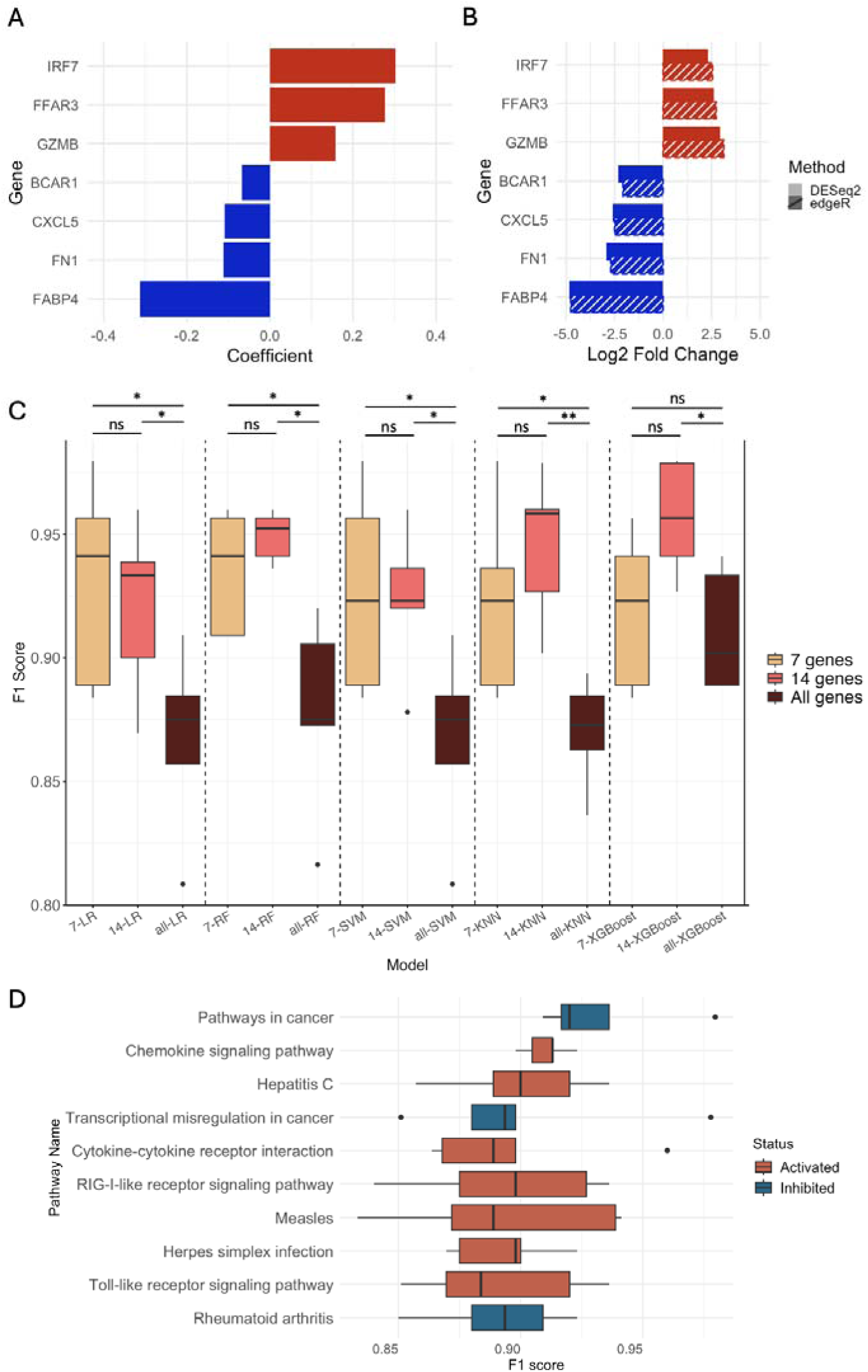
A) Regression coefficients of the 7 diagnostic markers selected from LASSO logistic regression model. B) Log_2_ fold change of the 7 diagnostic markers from differential expression analysis. C) Different models for classification of LRTI status. Boxplot shows F1 scores across 5-fold cross-validation. Significance levels of two-tailed Tuckey’s tests are indicated as follows: *P < 0.05, **P < 0.01, and “ns” for not significant. D) Top 10 LR classifiers using key genes in each pathway. Boxplot shows F1 scores across 5-fold cross-validation.

The seven-gene panel showed high diagnostic performance, with a median F1 score of 0.94 across logistic regression, random forest, and other classifiers, comparable to the 14-gene model (Figure 3C). Performance did not differ significantly between the two panels, and both clearly outperformed models trained on the full unfiltered gene set. Thus, the streamlined seven-gene panel offers accuracy comparable to the 14-gene model but with greater practicality.

Pathway analysis confirmed that LRTI-specific transcriptional changes clustered in immune and inflammatory signalling, notably chemokine, cytokine–receptor, and Toll-like receptor pathways (Figure 3D; Table 1).

**Table 1.**
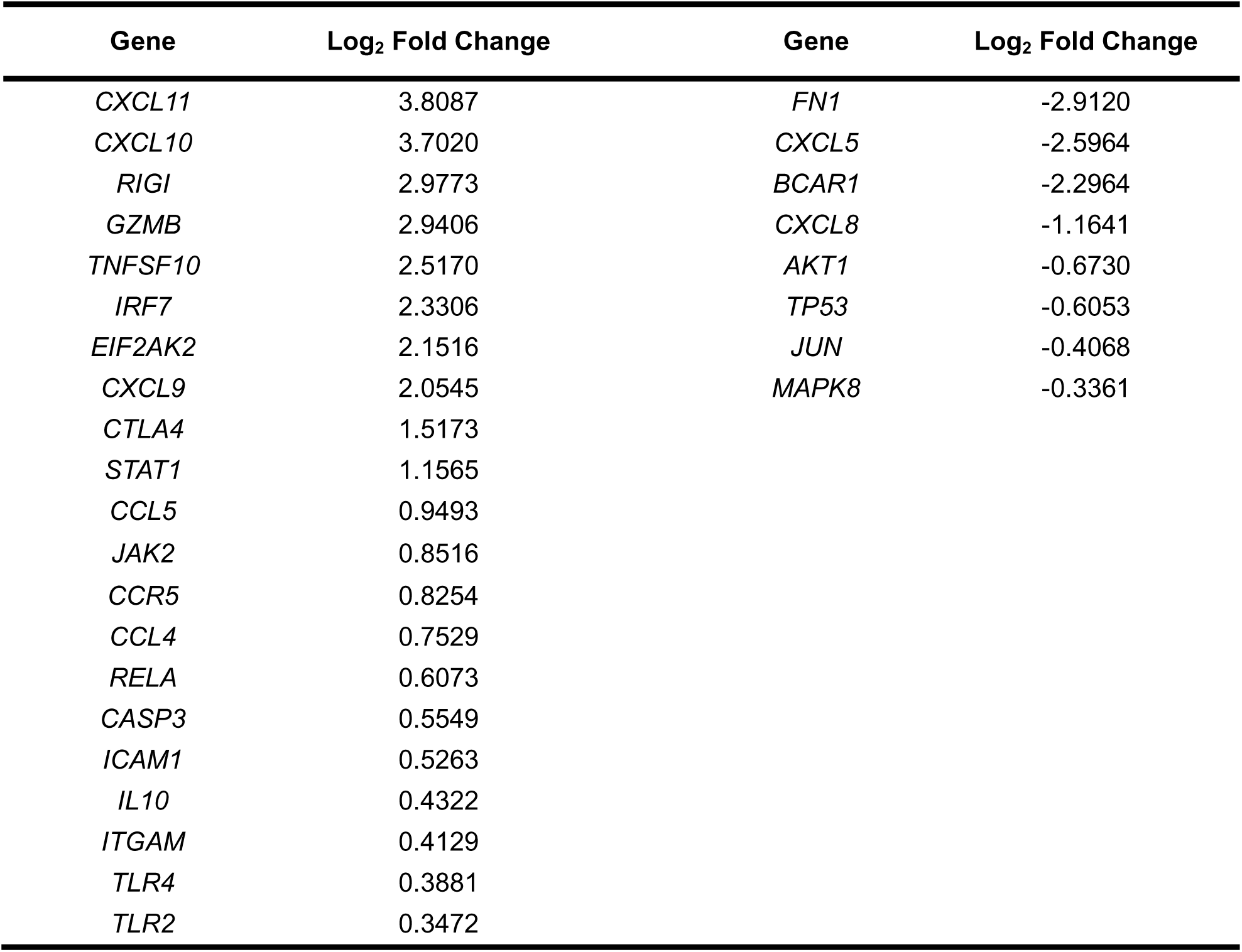
Key protein genes in the top 10 pathways. The table shows the gene symbols along with their log2 fold change values from DESeq2.

These results indicate that both discrete host gene signatures and broader immune pathways provide candidate biomarkers for early LRTI diagnosis in critically ill children.

### 3.4 Key Microbial Strains Associated with LRTI Progression

We next examined dominant microbial features associated with LRTI. To control for contaminants, microbial counts were adjusted against water controls. After filtering, viruses were detected in 91% of LRTI samples (107/117) versus 16% of controls (8/50), spanning 51 viral species. Differential abundance analysis identified respiratory syncytial virus (RSV) by both DESeq2 and edgeR, while rhinovirus A was detected only by DESeq2 (Figures 4A–B). RSV was therefore the sole virus consistently enriched in LRTI.

**Figure 4.**
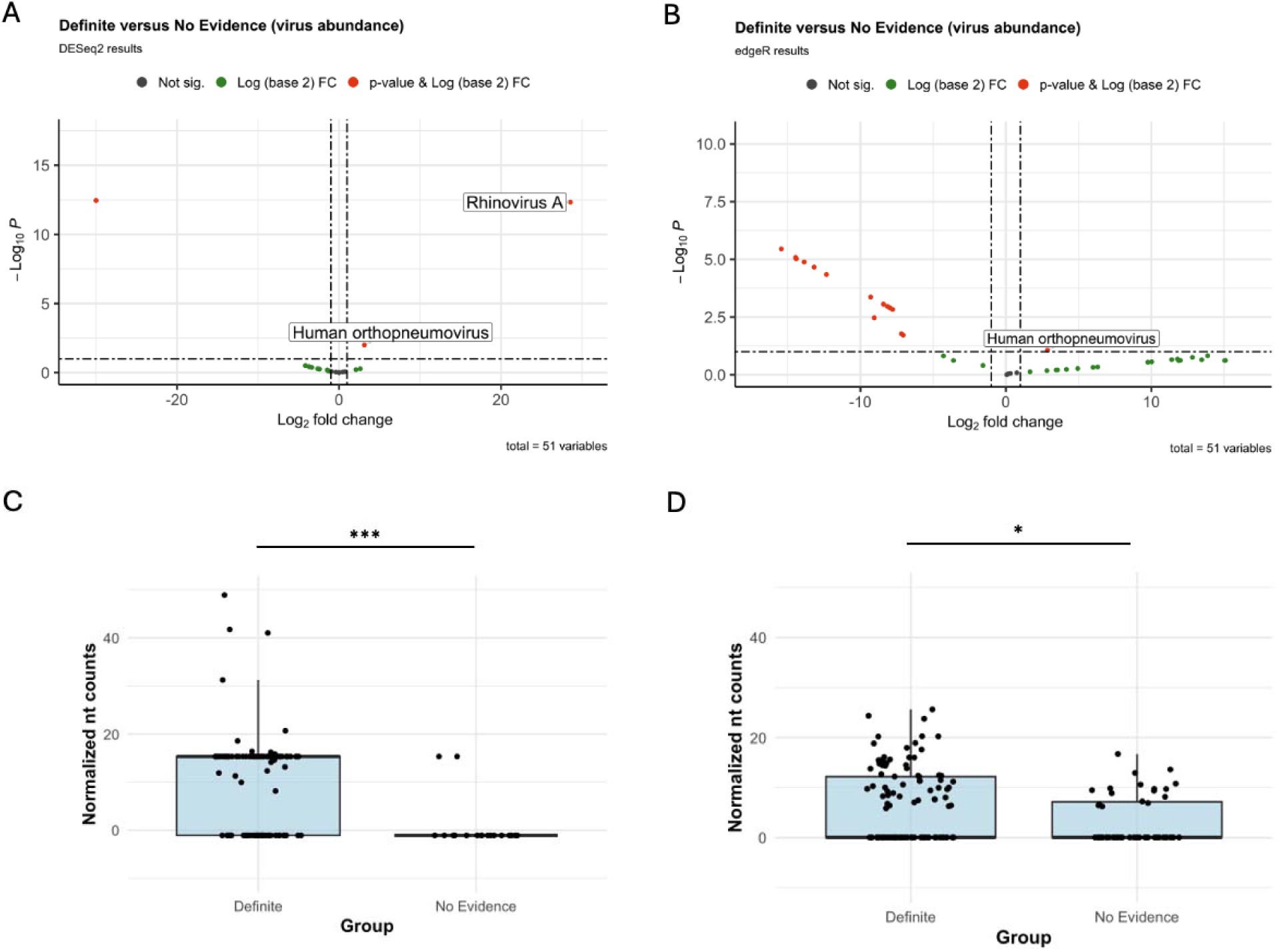
A) Volcano plot of differential analysis between LRTI and control cohorts on virus abundance using DESeq2. P-value < 0.1 and log_2_ fold change > 1 was deemed significant. B) Volcano plot on virus abundance using edgeR. Scatter bar chart representing abundance of C) Respiratory Syncytial Virus and D) Haemophilus influenzae between LRTI and control patients. Significance levels of two-tailed Welch’s two-sample t-test are indicated as follows: *P < 0.05, ***P < 0.001.

For bacteria and fungi, classification at the genus level showed Haemophilus and Moraxella enriched by both methods (SF5-6). At the species level, only *Haemophilus influenzae* was consistently identified across DESeq2 and edgeR (SF7-8). Subsequent analyses therefore focused on *H. influenzae*.

Quantitative comparisons using the Mann-Whitney U test confirmed RSV and *H. influenzae* enrichment in LRTI: RSV (P < 0.001, Figure 4C) and *H. influenzae* (P = 0.014, Figure 4D). Among LRTI samples, RSV was present in 67% (72/107) and *H. influenzae* in 49% (57/116).

Given RSV’s dominance, we explored its co-occurrence patterns with other microbial species using Kendall’s rank test. RSV co-occurred with five additional taxa (ST1), including *Trichoderma atroviride* and several bacterial species. The strong correlation between RSV and *H. influenzae* highlights a potential synergistic interaction exacerbating LRTI severity.

### 3.5 RSV-Associated Host Gene Expression and Network Perturbations Underpin LRTI Pathogenesis

Given RSV’s dominance, we assessed its impact on host responses through a meta-analysis of four independent transcriptomic studies of RSV-infected children (ST2). Using differential expression analysis, 111 genes were consistently identified as differentially expressed across at least two studies (Figure 5A), indicating conserved host responses to RSV.

**Figure 5.**
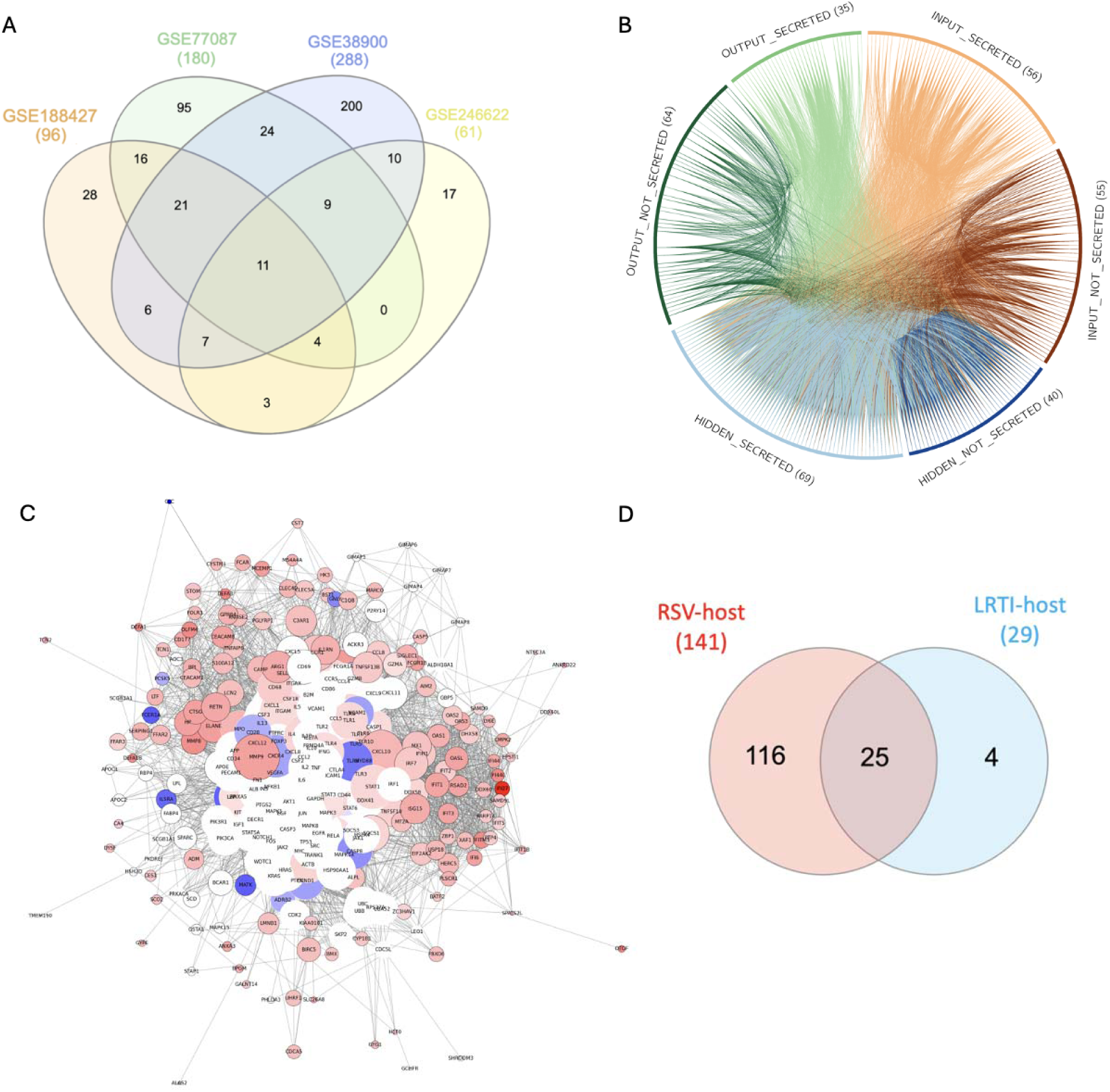
A) A Venn diagram showing the number of overlapping differentially expressed genes associated with RSV across four studies. B) A circus plot depicting interactions between input and output proteins through the PPI network. The plot includes input, output and key proteins, where 109 key proteins are in the hidden layer. The proteins are subdivided based on their secretion into bloodstream. The colored lines show PPI. C) A plot showing the network which includes only input and key proteins. Nodes with larger sizes have higher eigenvector centrality. The node colour reflects the log_2_ fold change in gene expression, with red indicating positive values, blue indicating negative values, and white indicating near-zero or N/A values. D) A Venn diagram showing key protein genes identified in the top 10 pathways and central in the RSV-host network.

Protein-protein interaction (PPI) network analysis showed that RSV-associated genes clustered in immune and inflammatory signalling pathways relevant to LRTI pathogenesis. Integrating RSV-specific DEGs with LRTI-associated DEGs produced an RSV–host interaction network containing 141 central proteins (Figures 5B–C). Many of these acted as intermediary “hidden layer” proteins linking RSV-induced responses to broader immune dysregulation.

Overlap analysis demonstrated that 25 of 29 key host proteins involved in the most significantly perturbed immune pathways in LRTI (Table 1) were also central in the RSV–host network (Figure 5D; ST3). This high degree of overlap highlights RSV as a primary driver of inflammatory signalling in paediatric LRTI.

Overall, these results suggest that RSV infection triggers conserved host gene networks that amplify immune activation and underpin LRTI progression.

### 3.6 Cytokine-Linked Key Network Proteins Underpin Bacterial LRTI Pathogenesis

We validated candidate biomarkers in an independent prospective cohort of 100 critically ill children with suspected LRTI (RASCALS). Of these, 78 patients were included in the analysis after excluding those without available samples or with insufficient remaining material following priority experiments. The median age was 1.2 years [IQR 0.4–5.2 years]. Mini-bronchoalveolar lavage (mini-BAL) and plasma blood samples were analysed alongside clinical outcomes, with ventilator-free days at 28 days serving as the primary severity marker.

Clinical data were reviewed by three PICU consultants to adjudicate LRTI status and classify infections as bacterial, non-bacterial, or non-LRTI. Children with bacterial LRTI (n=21) had fewer ventilator-free days compared to the non-bacterial group (n=57), suggesting greater disease severity and higher intervention needs, though the difference did not reach statistical significance (P = 0.10; Figure 6A). When the non-bacterial group was further subdivided into viral LRTI (n=39) and non-LRTI (n=18), bacterial LRTI again showed the poorest outcomes, although without statistical significance (P = 0.24, Figure 6B). These findings highlight the importance of characterising bacterial LRTI to enable timely diagnosis and intervention.

**Figure 6.**
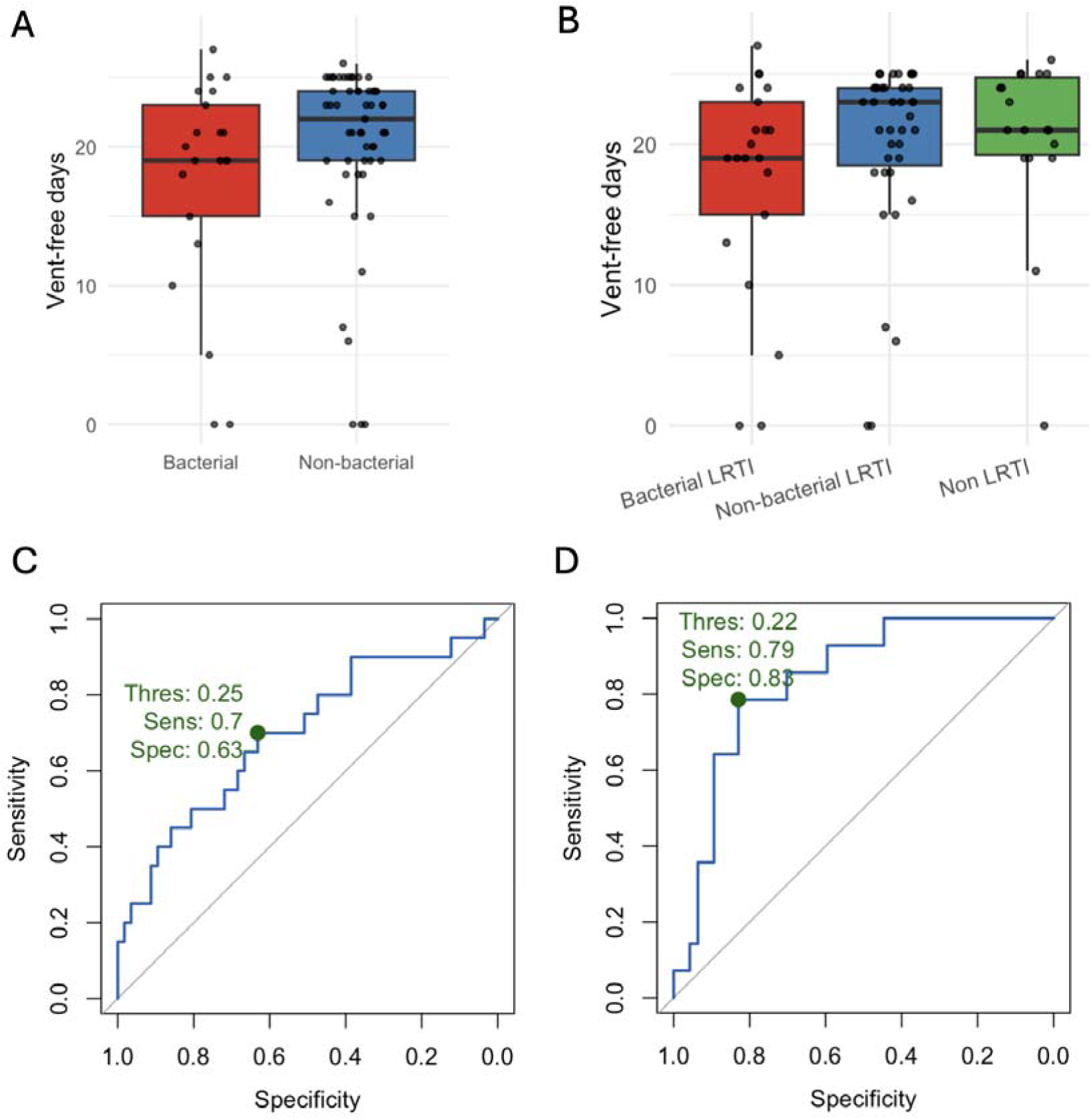
Distribution of ventilator-free days at 28 days following PICU admission A) between bacterial LRTI and non-bacterial groups; B) among bacterial LRTI, non-bacterial LRTI, and non LRTI groups. Receiver-operating characteristic (ROC) curves for logistic-regression model predicting LRTI status. C) Model using mini-BAL IL-1β + IL-4 + IL-8. B) Model using blood IL-6 + TRAIL. In each panel, the green dot marks the cut-off probability that yields the point on the curve closest to the top-left corner (optimal trade-off), with the corresponding threshold (Thres), sensitivity (Sens), and specificity (Spec) annotated. AUC, area under the ROC curve.

Respiratory syncytial virus (RSV) was detected in 11 patients, all of whom were diagnosed with LRTI, consistent with previous findings identifying RSV as a key risk factor. Of these, 10 were classified as viral LRTI and one as mixed infection.

To further delineate host responses specific to bacterial disease, we evaluated five cytokine markers identified through our network analysis: mini-BAL IL-1β (IL1B), IL-4, and IL-8 (CXCL8), and blood IL-6 and TRAIL (TNFSF10). Mini-BAL IL-1β, IL-4, and IL-8 levels were significantly elevated in bacterial LRTI compared to non-bacterial cases (P = 0.022, 0.027, and 0.019, respectively; Figure SF12, SF13). Similarly, plasma IL-6 levels were significantly higher in bacterial LRTI (P = 0.001), while TRAIL levels were significantly lower (P = 0.049; Figure SF12, SF13). Logistic regression models combining these cytokines achieved moderate-to-high diagnostic performance, with mini-BAL IL-1β + IL-4 + IL-8 achieving 65% diagnostic accuracy, (sensitivity 70%, specificity 63%; Figure 6C, Table 2) and blood IL-6 + TRAIL achieving 82% accuracy (sensitivity 79%, specificity 83%; Figure 6D, Table 3).

**Table 2.**
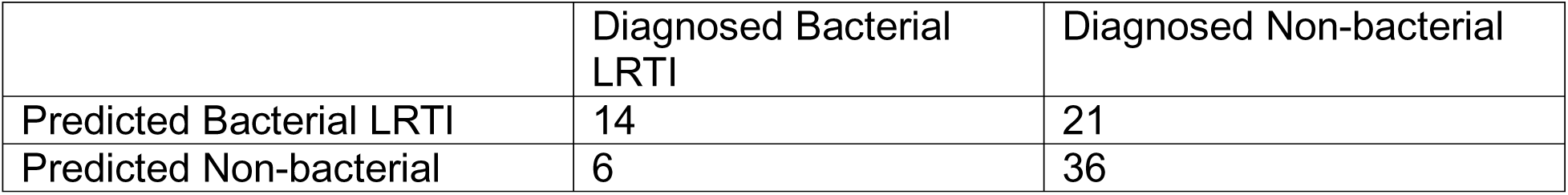
Mini-BAL IL-1β + IL-4 + IL-8 prediction versus clinical diagnosis of bacterial LRTI in the RASCALS cohort. 77 out of 78 patients have mini-BAL cytokine data.

|  | Diagnosed Bacterial LRTI | Diagnosed Non-bacterial |
| --- | --- | --- |
| Predicted Bacterial LRTI | 14 | 21 |
| Predicted Non-bacterial | 6 | 36 |

**Table 3.** Blood IL-6 + TRAIL prediction versus clinical diagnosis of LRTI in the RASCALS cohort. 61 out of 78 patients have blood cytokine data.

|  | Diagnosed Bacterial LRTI | Diagnosed Non-bacterial |
| --- | --- | --- |
| Predicted Bacterial LRTI | 11 | 8 |
| Predicted Non-bacterial | 3 | 39 |

Together, these results demonstrate that bacterial LRTI is associated with greater clinical severity, and that specific cytokine profiles can aid in distinguishing bacterial infections from non-bacterial causes in critically ill children.

## 4. Discussion

This study demonstrates that combining host and microbial biomarkers enables earlier and more accurate diagnosis of paediatric LRTI in critically ill children. By integrating host transcriptomic and airway microbial data, we identified RSV, non-typeable Haemophilus influenzae, and a seven-gene host signature as key contributors to LRTI. Importantly, validation in the prospective RASCALS cohort showed that host cytokines achieved diagnostic accuracies of 65% (mini-BAL) and 82% (blood) for bacterial LRTI and correlated with clinical outcomes.

These findings provide a proof-of-concept for multi-omic diagnostics in PICU.

The seven-gene host panel (IRF7, FFAR3, GZMB, FABP4, FN1, CXCL5, BCAR1) demonstrated strong diagnostic accuracy and biological relevance. IRF7, GZMB, and CXCL5 are established mediators of antiviral immunity,^21–23^ while FFAR3 and FABP4 regulate macrophage responses,^24,25^ and FN1 and BCAR1 contribute to tissue homeostasis.^26,27^ Four of these genes also overlapped with RSV-driven dysregulation, highlighting the central role of RSV in shaping host responses. The predominance of RSV and NTHi as microbial markers is consistent with their established role in paediatric LRTI.^28,29^

Pathway analysis confirmed broad activation of antiviral and inflammatory signalling, including chemokine, cytokine–receptor, RIG-I-like receptor, and Toll-like receptor pathways. Interestingly, pathways linked to cancer and autoimmune conditions were downregulated, suggesting a physiological shift prioritising acute antiviral defence. These findings support the concept that host immune activation, rather than pathogen features alone, drives disease progression.

RSV also predisposes to bacterial co-infection. Mechanistic studies show RSV can impair neutrophil function and promote bacterial adhesion to respiratory epithelial cells,^30^ and we observed enrichment of Haemophilus and Streptococcus in RSV-associated LRTI. Although causality cannot be established, our integration analysis identified 25 genes linking RSV-triggered responses with secondary bacterial pathogenesis, providing new molecular insights.

Clinically, these biomarkers could be translated into PCR- or ELISA-based assays using samples already collected in ventilated children. Early detection of RSV, H. influenzae, and key host markers could enable rapid bedside diagnostics, improve risk stratification, and inform antimicrobial decisions.

This study has limitations. Reliance on mini-BAL specimens may limit generalisability to blood-based diagnostics. Functional validation in experimental models is required to confirm mechanistic inferences. Taxonomic classification at the genus level risks obscuring pathogen-specific signals, and exclusion of RSV and H. influenzae counts from classifiers due to dataset imbalance constrains predictive modelling. Future studies should address these limitations, ideally in multi-centre cohorts with larger sample sizes.

## 5. Conclusion

This study shows that integrating host and microbial markers enables early and accurate diagnosis of paediatric LRTI in critically ill children. A seven-gene host panel, together with RSV, *Haemophilus influenzae*, and cytokine markers (IL-1β, IL-4, IL-8, IL-6, and TRAIL), achieved strong diagnostic performance and correlated with clinical outcomes.

These findings support development of PCR- or ELISA-based bedside assays for rapid detection in ventilated patients, with potential to improve risk stratification and antimicrobial stewardship. Multi-centre validation and functional studies are now needed to translate these biomarkers into routine clinical use.

## Supporting information

SF1-SF13, Table S1-S9, List 1-3

## Data Availability

All data, code, and software packages used, including versions and citations, are available at https://github.com/zongtaiwu/mphil-paediatric-LRTI. The raw data, including patient adjudication, host gene counts, and microbial taxon counts were retrieved from Mick et al.10 at https://github.com/eranmick/pediatric-mNGS-LRTI-classifier. The code for protein network analysis was adapted based on the code from Han et al.20 at https://github.com/wchwang/COVID19.

https://github.com/wchwang/COVID19

## Acknowledgements

We thank Dr Constantinos Kanaris, Dr Shruti Agrawal, and Dr David Inwald for their contribution to the clinical adjudication process. Final diagnoses of LRTI or non-LRTI were assigned at hospital discharge and adjudicated under their supervision.

