## Supplementary material for "Developing an Early Diagnostic Signature and Deciphering the Microbial-Host Dynamics in Lower Respiratory Tract Infection (LRTI) in Paediatric Intensive Care Unit (PICU) Patients": SF1-SF13, Table S1-S9, List 1-3

**SF1. *Flow chart of the analytical framework utilised in this study.***

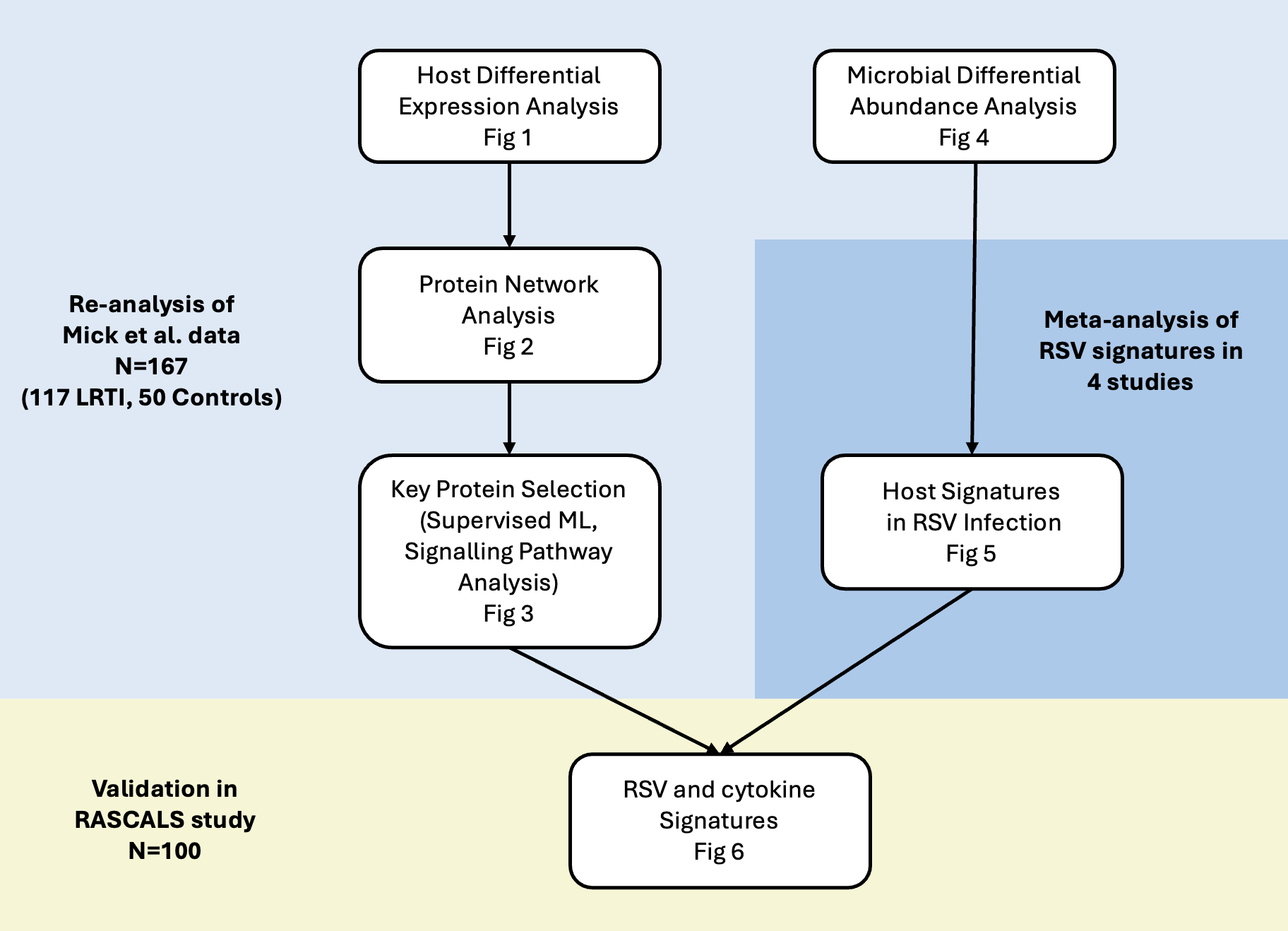

**SF2.** Plot showing all nodes and edges in the full LRTI network including input proteins and all hidden layer proteins. Nodes with larger sizes have higher eigenvector centrality. The node colour reflects the log_2_ fold change in gene expression, with red indicating positive values, blue indicating negative values, and white indicating near-zero or N/A values.

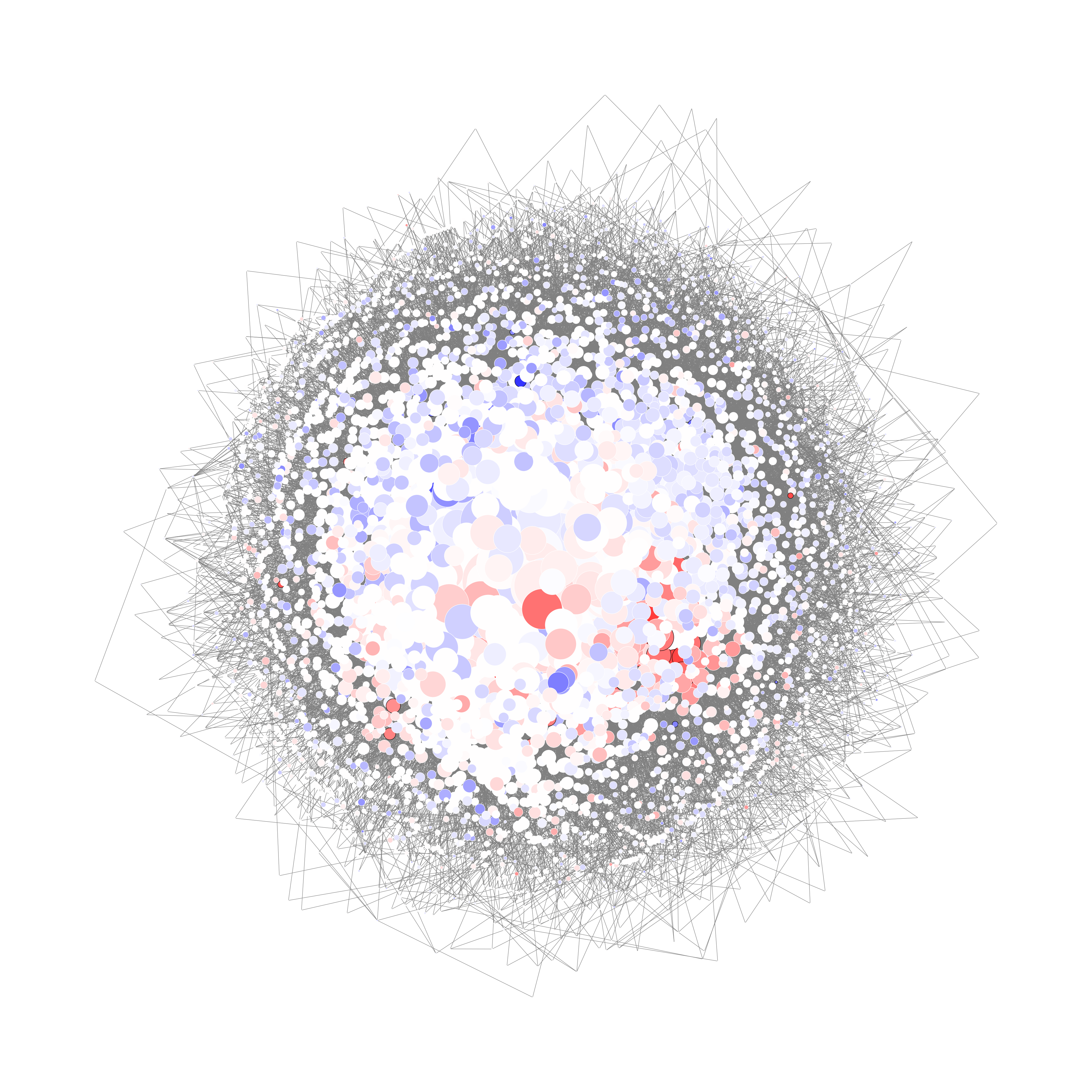

**SF3.** Plot showing the subnetwork which includes all edges containing key or input proteins. Nodes with larger sizes have higher eigenvector centrality. The node colour reflects the log_2_ fold change in gene expression, with red indicating positive values, blue indicating negative values, and white indicating near-zero or N/A values.

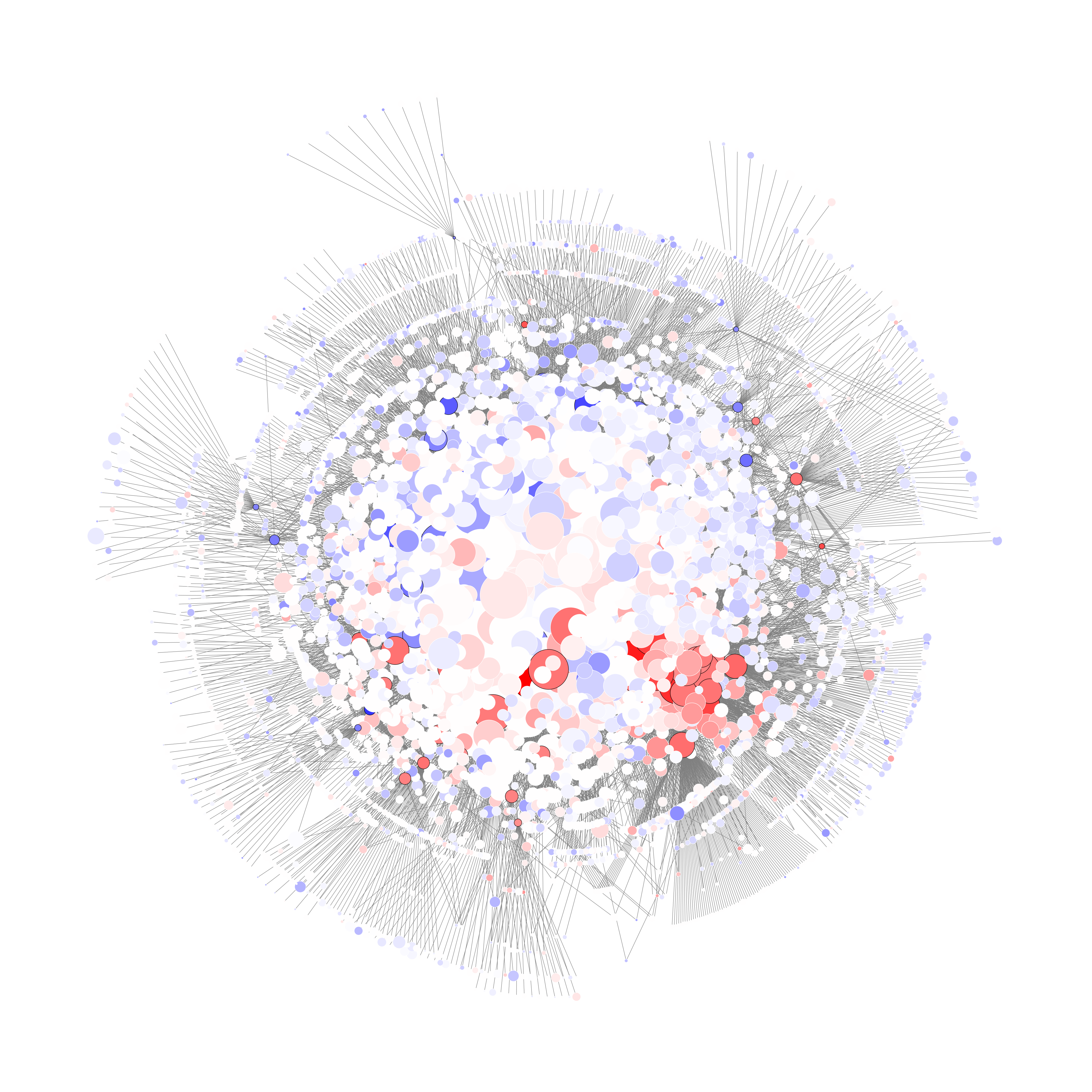

**SF4.** All LR classifiers using key genes in each pathway. Pathways in descending order of the mean F1 score across 5 folds.

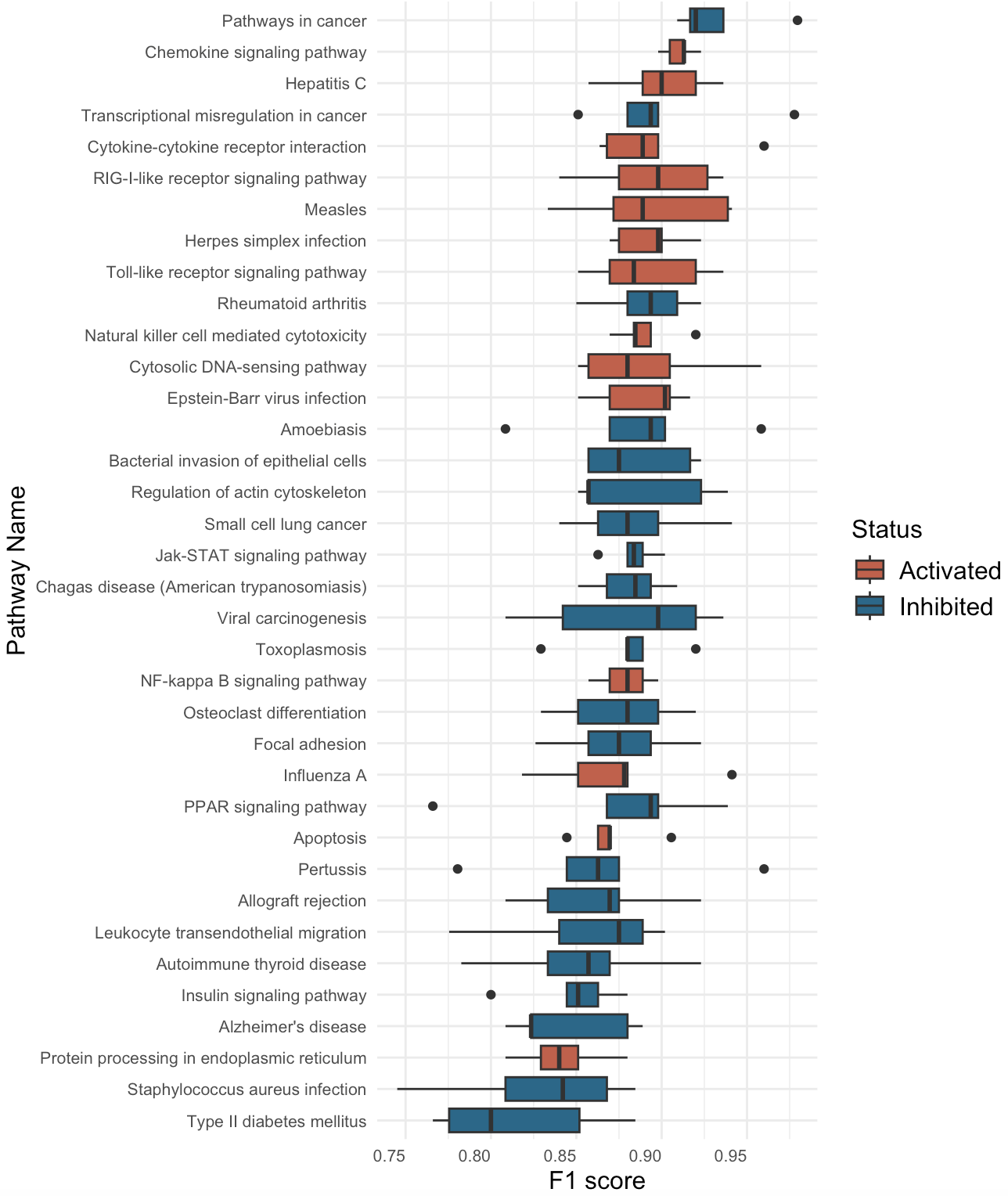

**SF5.** Volcano plot on bacteria/fungi grouped by genus using DESeq2.

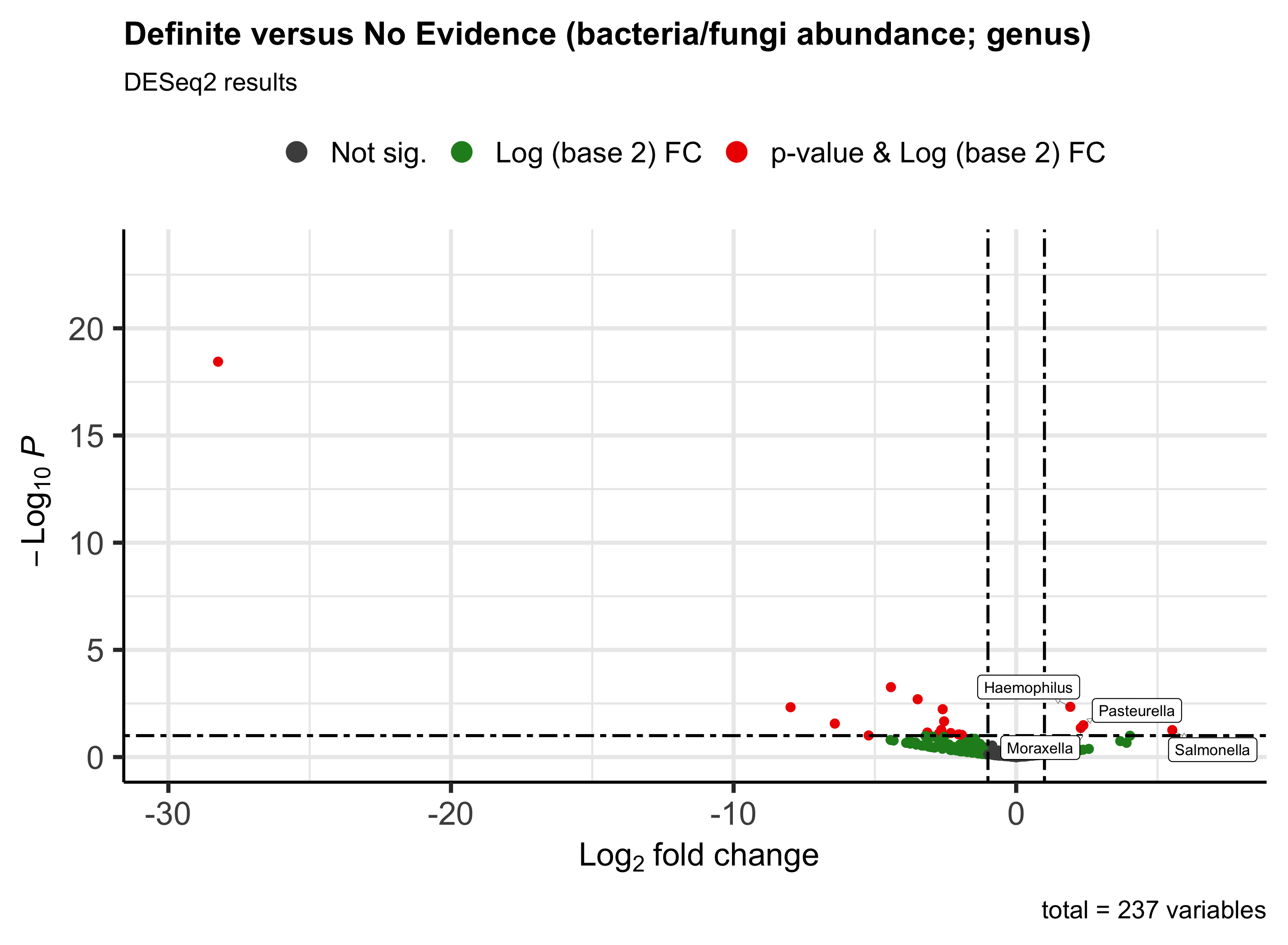

**SF6.** Volcano plot on bacteria/fungi grouped by genus using edgeR.
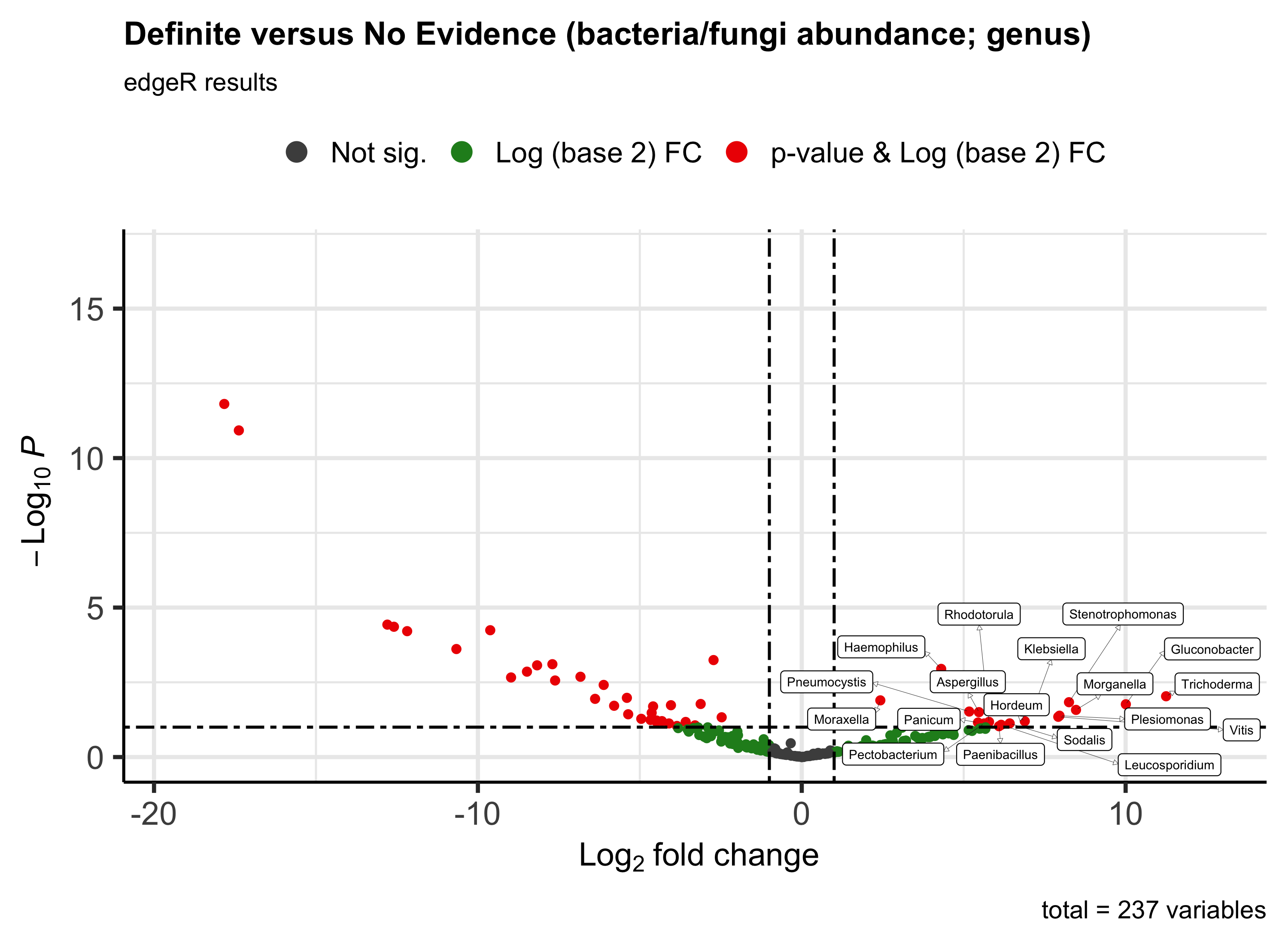

**SF7.** Volcano plot on bacteria/fungi grouped by species using DESeq2.

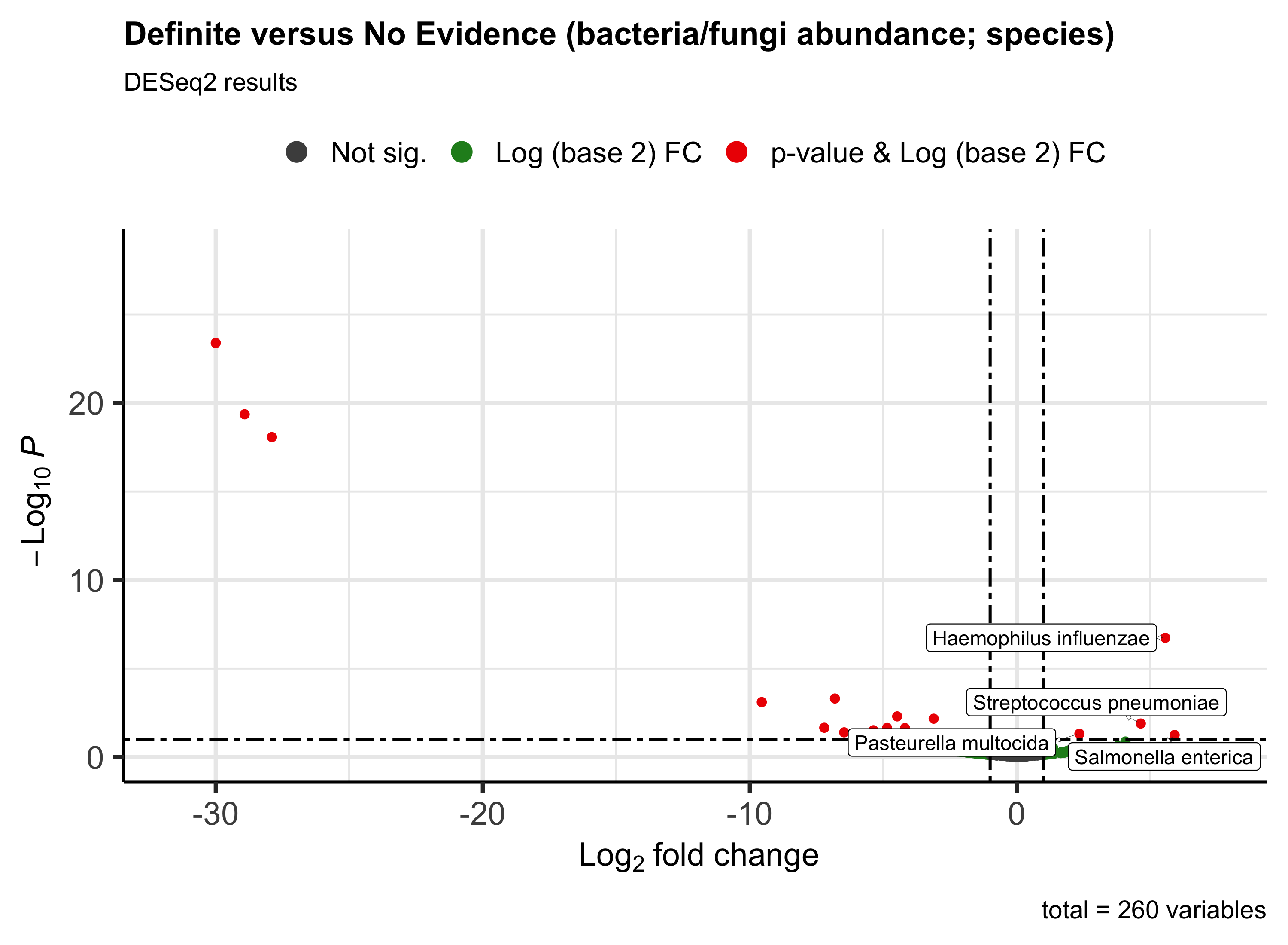

**SF8.** Volcano plot on bacteria/fungi grouped by species using edgeR.

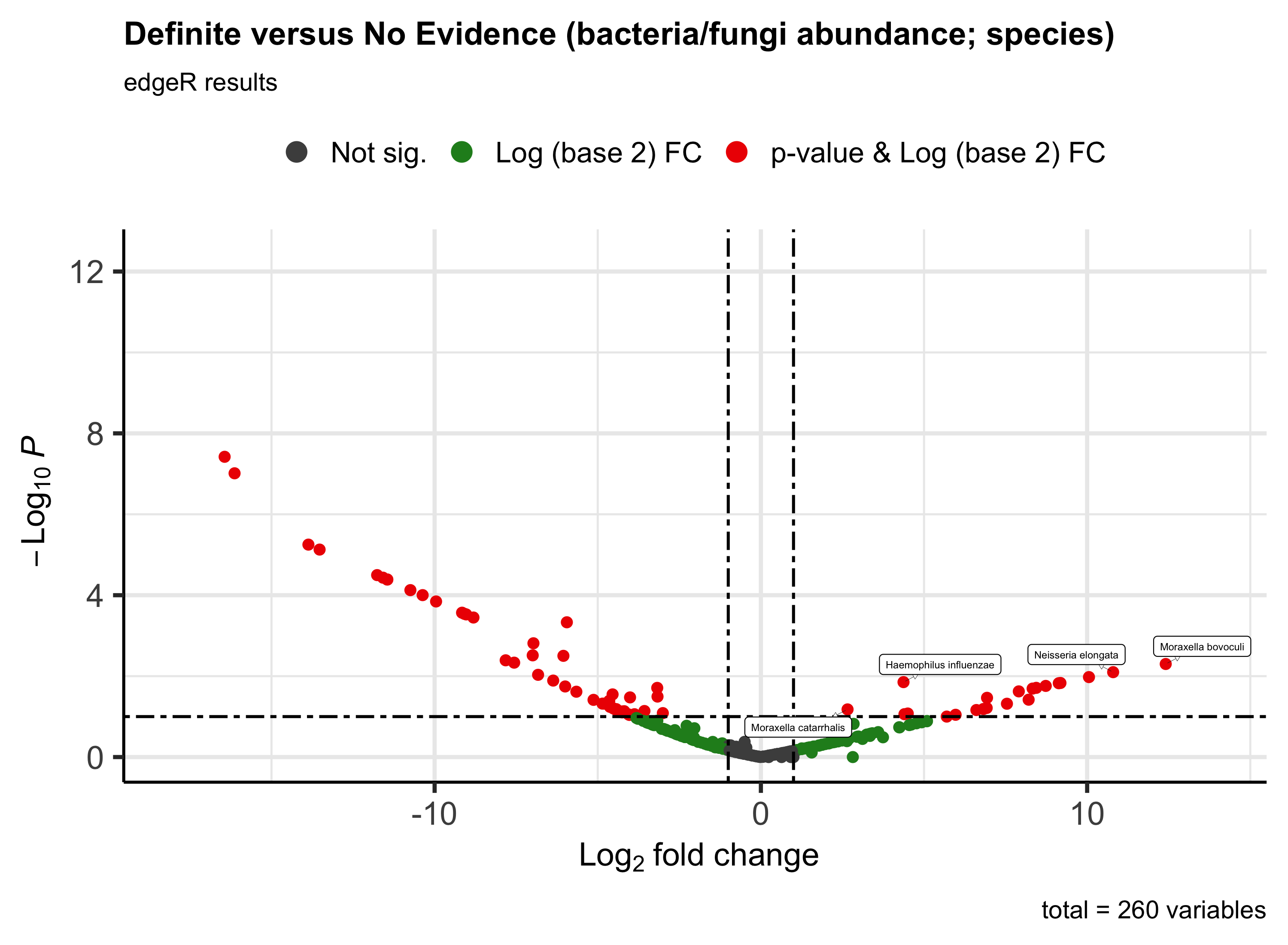

**SF9.** Plot showing all nodes and edges in the full RSV-host network including input, output, and all hidden layer proteins. Nodes with larger sizes have higher eigenvector centrality. The node colour reflects the log_2_ fold change in gene expression, with red indicating positive values, blue indicating negative values, and white indicating near-zero or N/A values.

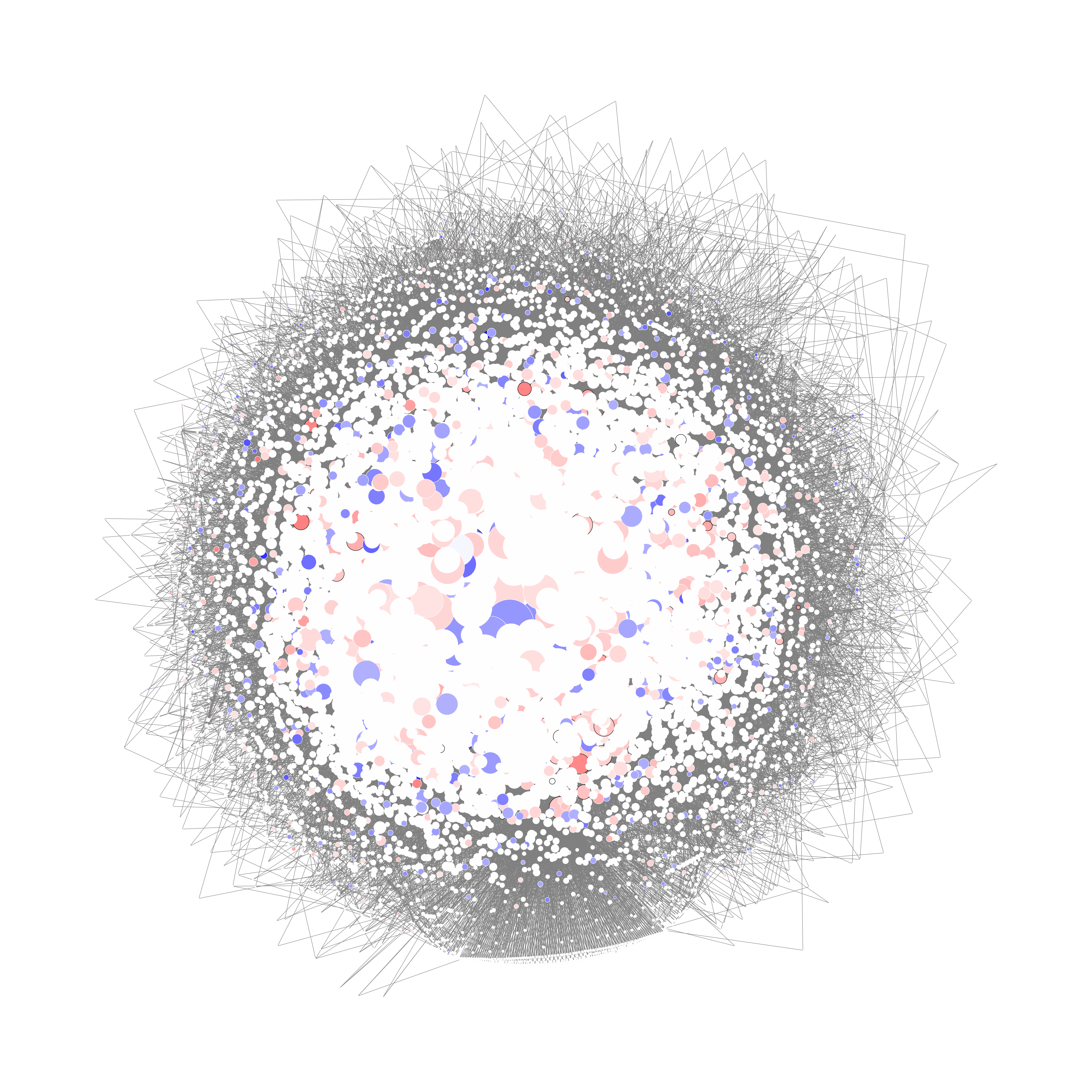

**SF10.** Plot showing the subnetwork which includes all edges containing key, input and output proteins. Nodes with larger sizes have higher eigenvector centrality. The node colour reflects the log_2_ fold change in gene expression, with red indicating positive values, blue indicating negative values, and white indicating near-zero or N/A values.

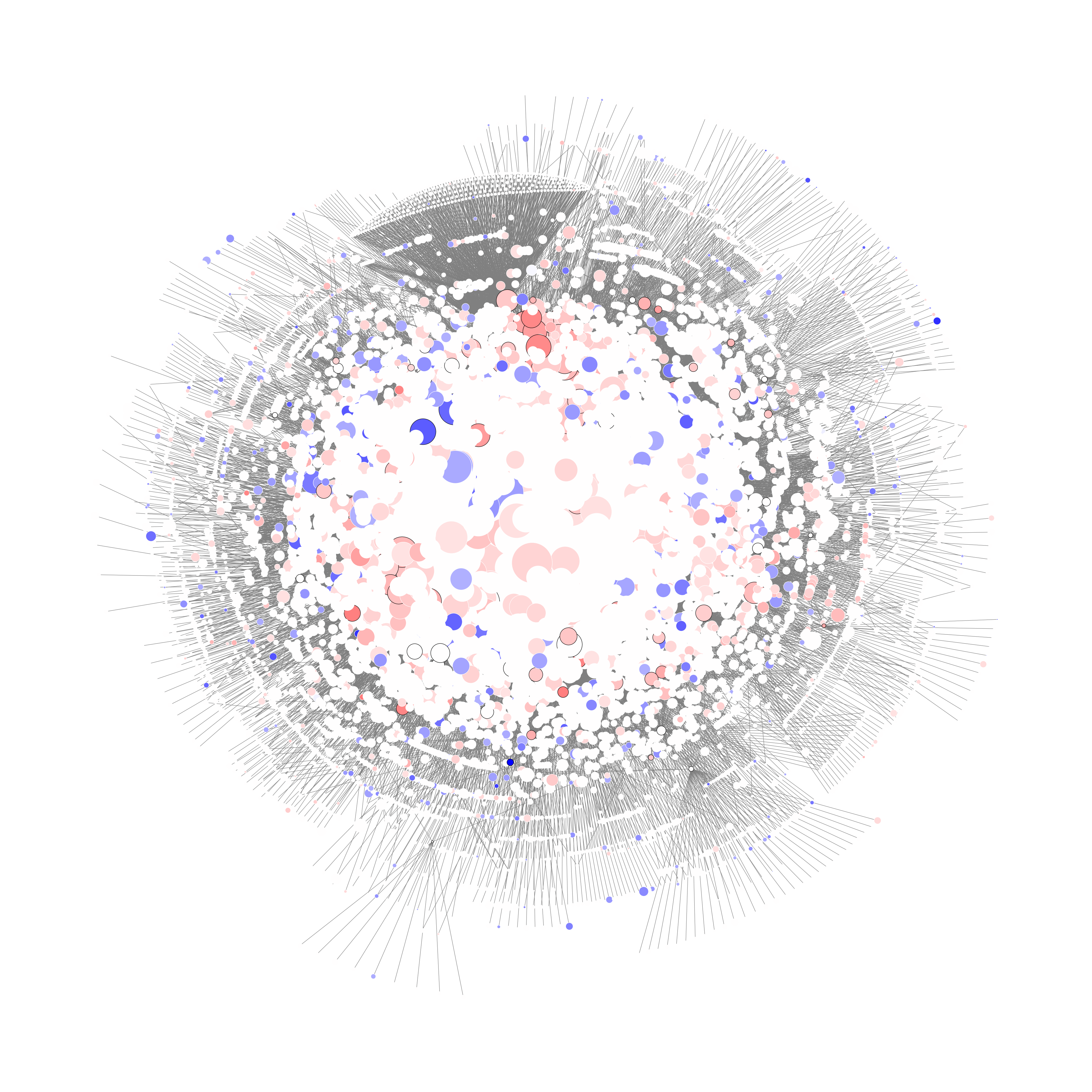

**SF11.** Graphic illustration of the nested 5-fold cross-validation method. The best-performing model from the inner loop is used to train the train fold in the outer loop.

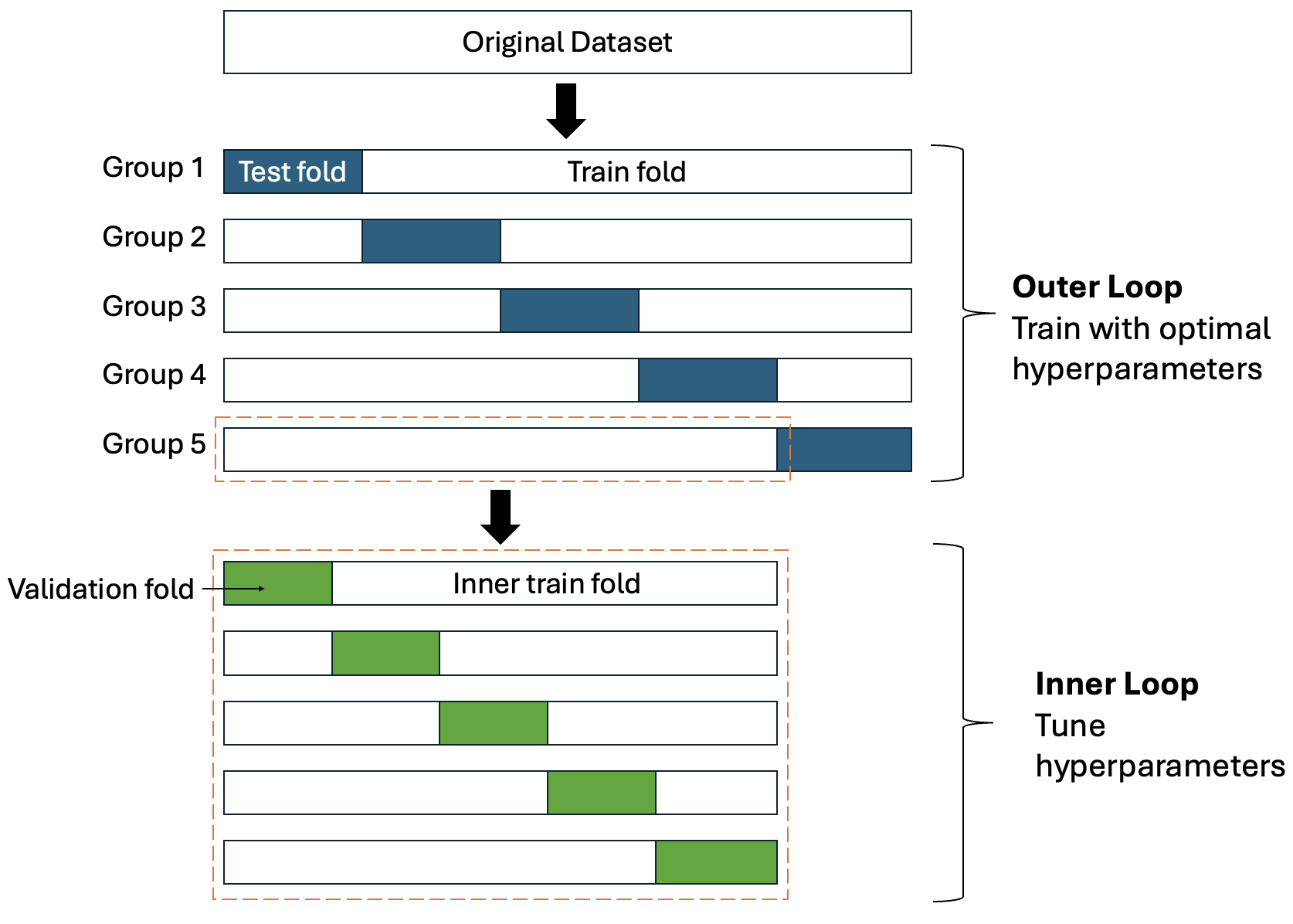

**SF12.** A) to D): Comparison of bronchoalveolar-lavage level of A) IL-8, B) IL-4, C) TRAIL, and D) IL-4:TRAIL ratio among RSV(+), RSV(-) LRTI and RSV(-) non-LRTI patients. G) to H): Comparison of blood level of A) IL-8, B) IL-4, C) TRAIL, and D) IL-4:TRAIL ratio among RSV(+), RSV(-) LRTI and RSV(-) non-LRTI patients.

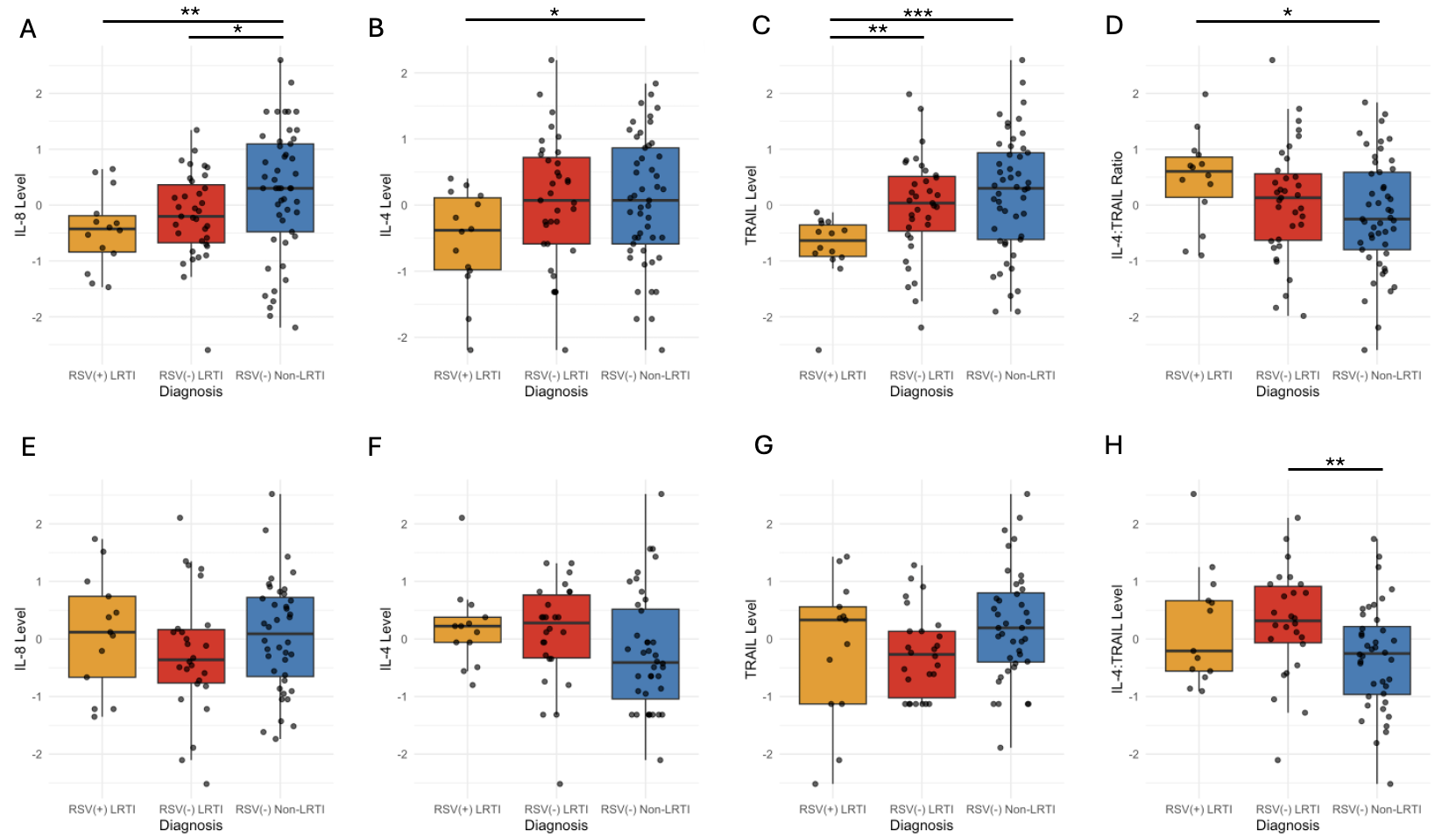

**SF13.** Receiver-operating characteristic (ROC) curves for logistic-regression model predicting LRTI status. A) Model using bronchoalveolar-lavage IL-8. B) Model using blood IL-4∶TRAIL ratio. In each panel, the green dot marks the cut-off probability that yields the point on the curve closest to the top-left corner (optimal trade-off), with the corresponding threshold (Thres), sensitivity (Sens), and specificity (Spec) annotated. AUC, area under the ROC curve.

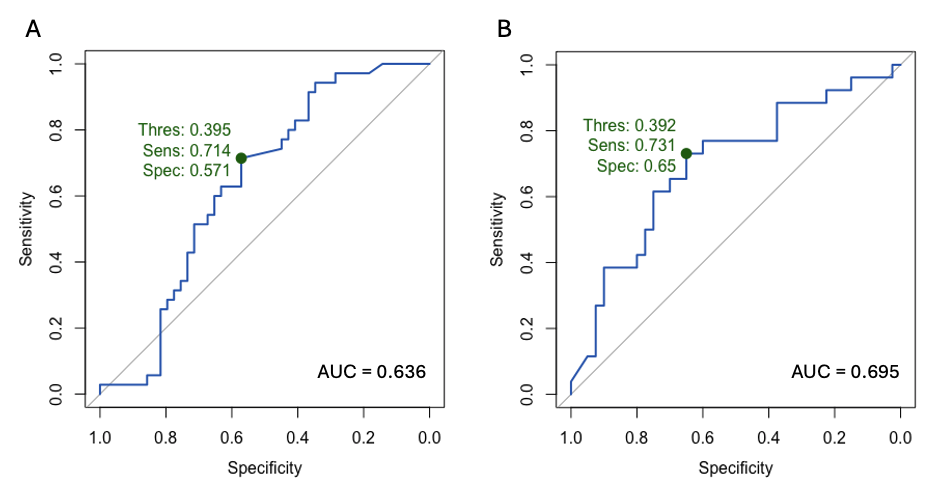

**Table S1.** Bacteria/Fungi species correlated with RSV. The correlation was tested using Kendall rank correlation coefficient and p-value < 0.1 was deemed significant.

| **Species** |  | **Correlation** | **P Value** |
| --- | --- | --- | --- |
| *Trichoderma atroviride* |  | 0.16341 | 0.04658 |
| *Enterocloster bolteae* |  | 0.14638 | 0.07404 |
| *Streptococcus pseudopneumoniae* |  | 0.14657 | 0.07483 |
| *Haemophilus influenzae* |  | 0.12221 | 0.09569 |
| *Staphylococcus capitis* |  | 0.13691 | 0.09610 |

**Table S2.** Description of datasets used in the meta-analysis.

| **GEO accession** | **Authors** | **Year** | **Country** | **Selected Patients** | **Platform** | **Total Sample**  **(No.)** | **RSV (No., %)** | **Control (No., %)** |
| --- | --- | --- | --- | --- | --- | --- | --- | --- |
| GSE188427 | Thys et al. | 2022 | Belgium | Children <2 years old with RSV infection at presentation and age-matched healthy controls | GPL25336 (Affymetrix Clariom S Pico Assay HT) | 353 | 147, 41.6% | 51,  14.4% |
| GSE77087 | de Steenhuijsen Piters et al. | 2016 | USA | Children with RSV disease and healthy matched controls | GPL10558 (Illumina HumanHT-12 V4.0 expression beadchip) | 104 | 81, 77.9% | 23,  22.1% |
| GSE38900 | Meijas et al. | 2013 | USA | Children < 2 years old with RSV and healthy matched controls | GPL6884 (Illumina HumanWG-6 V3.0 expression beadchip) | 205 | 107, 52.2% | 31, 15.1% |
| GSE246622 | Zivanovic et al. | 2023 | Belgium | Children < 12 months old with severe RSV  and healthy matched controls | GPL31262 (Affymetrix human GO_Screen_hu Assay) | 533 | 96, 18.0% | 56, 10.5% |

**Table S3.** Key protein genes in the top 10 pathways and central in the RSV-host network (names in bold). The table shows the gene symbols along with their log_2_ fold change values from DESeq2.

| **Gene** | **Log_2_ Fold Change** |  | **Gene** | **Log_2_ Fold Change** |
| --- | --- | --- | --- | --- |
| **CXCL10** | 3.7020 |  | **FN1** | -2.9120 |
| **RIGI** | 2.9773 |  | **BCAR1** | -2.2964 |
| **GZMB** | 2.9406 |  | **CXCL8** | -1.1641 |
| **IRF7** | 2.3306 |  | **AKT1** | -0.6730 |
| **CXCL9** | 2.0545 |  | **TP53** | -0.6053 |
| **CTLA4** | 1.5173 |  | **JUN** | -0.4068 |
| **STAT1** | 1.1565 |  | **MAPK8** | -0.3361 |
| **CCL5** | 0.9493 |  |  |  |
| **JAK2** | 0.8516 |  |  |  |
| **CCR5** | 0.8254 |  |  |  |
| **CCL4** | 0.7529 |  |  |  |
| **RELA** | 0.6073 |  |  |  |
| **CASP3** | 0.5549 |  |  |  |
| **ICAM1** | 0.5263 |  |  |  |
| **IL10** | 0.4322 |  |  |  |
| **ITGAM** | 0.4129 |  |  |  |
| **TLR4** | 0.3881 |  |  |  |
| **TLR2** | 0.3472 |  |  |  |

**Table S4.** RSV‐based prediction versus clinical diagnosis of LRTI in the RASCALS cohort (N=100).

|  | Diagnosed LRTI | Non-LRTI |
| --- | --- | --- |
| RSV-predicted LRTI | 14 | 0 |
| RSV-predicted Non-LRTI | 33 | 53 |

**Table S5.** BAL-IL-8‐based prediction versus clinical diagnosis of LRTI in the RSV-negative patents with BAL sample analysed (N=84).

|  | Diagnosed LRTI | Non-LRTI |
| --- | --- | --- |
| BAL-IL-8-predicted LRTI | 25 | 21 |
| BAL-IL-8-predicted Non-LRTI | 10 | 28 |

**Table S6.** Blood-IL-4:TRAIL-ratio‐based prediction versus clinical diagnosis of LRTI in the RSV-negative patents with blood sample analysed (N=66).

|  | Diagnosed LRTI | Non-LRTI |
| --- | --- | --- |
| Blood-ratio-predicted LRTI | 19 | 14 |
| Blood-ratio-predicted Non-LRTI | 7 | 26 |

**Table S7. Plasma cytokine concentrations in bacterial versus non-bacterial LRTI (RASCALS cohort).**
Median cytokine concentrations (pg/mL) with interquartile ranges are shown for children adjudicated as bacterial (n=20) or non-bacterial (n=58) LRTI. Comparisons were performed using the Mann–Whitney U test with Benjamini–Hochberg correction for multiple comparisons. IL-6 and CRP showed significant differences between groups, whereas other plasma cytokines did not.

| **Variable** | **Total** | **Bacterial LRTI present** | **No bacterial LRTI present** | **p-value** |
| --- | --- | --- | --- | --- |
|  | **N = 78** | **N = 20** | **N = 58** |  |
| **Demographics** |  |  |  |  |
| Age (years) - median (IQR) | 1.2 (0.4 – 5.2) | 4.0 (1.0 – 13.2) | 1.0 (0.3 – 4.4) | 0.046 |
| Sex (male) - n (%) | 41 (53) | 9 (45) | 32 (55) | 0.297 |
| Weight (kilograms) - median (IQR) | 10.9 (5.8 – 18.2) | 16.0 (7.9 – 40.4) | 9.6 (5.3 – 14.4) | 0.053 |
| Significant co-morbidity - n (%) | 25 (32) | 8 (40) | 17 (29) | 0.488 |
| PIM3 score - median (IQR) | 3.2 (0.5 – 4.8) | 3.6 (1.8 – 6.5) | 1.3 (0.5 – 9.4) | 0.034 |
| **Primary diagnostic category** - n (%) |  |  |  |  |
| Respiratory | 50 (64) | 10 (50) | 40 (69) | 0.177 |
| Neurological | 6 (8) | 3 (15) | 3 (5) | 0.172 |
| Cardiovascular | 7 (9) | 1 (5) | 6 (10) | 0.671 |
| Trauma | 2 (3) | 2 (10) | 0 (0) | 0.063 |
| Post-operative care | 5 (6) | 2 (10) | 3 (5) | 0.598 |
| Other |  |  |  |  |
| **Admission source** - n (%) |  |  |  |  |
| Retrieval team | 54 (69) | 14 (70) | 40 (69) | 1.000 |
| Emergency department | 9 (12) | 3 (15) | 6 (10) | 0.687 |
| Paediatric ward | 9 (12) | 2 (10) | 7 (12) | 1.000 |
| Theatre and recovery | 6 (8) | 2 (10) | 4 (7) | 0.643 |
| **Clinical factors at time of sampling** |  |  |  |  |
| Observations – mean (SD) |  |  |  |  |
| Temperature (^°^C) | 37.0 (1.0) | 37.3 (1.1) | 36.9 (0.9) | 0.082 |
| Fraction of inspired oxygen (%) | 44.5 (19.6) | 37.0 (16.3) | 47.3 (20.1) | 0.025 |
| Mean airway pressure (cm H_2_O) | 12.1 (3.1) | 11.2 (2.7) | 12.4 (3.2) | 0.098 |
| Secretions - n (%)^*^ |  |  |  |  |
| Absent/minimal | 20 (26) | 12 (60) | 33 (58) | 0.887 |
| Present non-purulent | 45 (58) | 6 (30) | 6 (11) | 0.054 |
| Present purulent | 12 (15) | 3 (15) | 17 (30) | 0.152 |
| Chest x-ray - n (%)^*^ |  |  |  |  |
| Diffuse/patchy infiltrate | 25 (32) | 5 (26) | 20 (34) | 0.407 |
| Localised infiltrate | 21 (27) | 7 (37) | 14 (24) | 0.367 |
| No infiltrate | 19 (25) | 6 (32) | 13 (22) | 0.521 |
| Other | 12 (16) | 2 (11) | 10 (17) | 0.424 |
| Haematology and biochemistry - median (IQR) | | | | |
| White cell count (10^9^/L) | 8.8 (6.0 – 14.7) | 8.9 (6.9 – 14.2) | 8.8 (5.8 – 15.3) | 0.711 |
| Neutrophils (10^9^/L) | 4.8 (3.2 – 10.7) | 6.1 (4.1 – 10.7) | 4.7 (2.9 – 11.3) | 0.432 |
| Lymphocytes (10^9^/L) | 2.1 (1.1 – 3.1) | 1.9 (0.9 – 3.0) | 2.2 (1.2 – 3.4) | 0.405 |
| Platelets (10^9^/L) | 281 (217 – 411) | 269 (191 – 334) | 306 (235 – 416) | 0.340 |
| C-reactive protein (mg/L)^$^ | 28.0 (7.0 – 88.5) | 90.0 (39.0 – 257.5) | 17.0 (4.5 – 55.0) | <0.001 |
| **Outcomes** |  |  |  |  |
| Days free of treatment at 28 days – mean (SD) |  |  |  |  |
| Antimicrobial therapy | 20.4 (6.8) | 16.5 (8.8) | 21.8 (5.3) | 0.026 |
| Mechanical ventilation | 19.7 (6.5) | 20.5 (5.8) | 17.5 (7.8) | 0.182 |
| Inotropes | 26.2 (5.0) | 26.9 (3.5) | 24.3 (7.5) | 0.139 |
| PICU admission | 17.2 (7.4) | 18.1 (6.7) | 14.8 (8.6) | 0.167 |
| Survival to hospital discharge - n (%) | 74 (95) | 19 (95) | 55 (95) | 0.285 |
| ^a = Mann-Whitney U test; b = Chi-square test for independence; c = Student’s t-test for equality of means, two-sided p, equal variance not assumed; d = Fishers exact test; CAP = community acquired pneumonia; HAP: hospital acquired pneumonia; IQR = Interquartile range; LRTI = Lower respiratory tract infection; PICU = Paediatric intensive care unit; SD = Standard deviation; VAP: ventilator associated pneumonia; * data missing for one patient in the medical record. $ test not undertaken in 3 patients with bacterial LRTI and 10 patients without bacterial LRTI.^ | | | | |

**No cytokine reached statistical significance between groups.**

**Table S8. Mini-BAL cytokine concentrations in bacterial versus non-bacterial LRTI (RASCALS cohort).**
Median cytokine concentrations (pg/mL) with interquartile ranges are shown for children with bacterial (n=20) or non-bacterial (n=58) LRTI. Comparisons were performed using the Mann–Whitney U test with Benjamini-Hochberg correction. No cytokine reached statistical significance between groups.

| Cytokine (pg/mL) | Bacterial LRTI  N = 20 | No bacterial LRTI  N = 58 | p-value* |
| --- | --- | --- | --- |
| G-CSF | 296.4 (12.3 – 1914.9) | 74.6 (31.5 – 217.9) | 0.460 |
| IL-1β | 7.2 (3.5 – 33.0) | 4.3 (1.7 – 11.2) | 0.568 |
| IL-6 | 60.6 (18.8 – 767.3) | 14.5 (2.9 – 36.6) | 0.040 |
| IL-8 | 44.5 (14.0 – 245.0) | 26.4 (11.3 – 62.1) | 0.362 |
| IP-10 | 1065.9 (338.8 – 2531.4) | 580.0 (267.9 -1908.9) | 0.436 |
| MCP | 43.5 (18.3 – 120.9) | 25.5 (12.5 – 79.1) | 0.333 |
| MIP-1α | 3.5 (1.9 – 5.7) | 2.2 (1.3 – 4.9) | 0.386 |
| TRAIL | 11.3 (5.8 – 31.8) | 21.8 (11.3 – 57.6) | 0.096 |
| ^*Mann-Whitney U test with Benjamani and Hochberg correction for multiple comparisons^ | | | |

| Cytokine (pg/mL) | Bacterial LRTI  N = 20 | No bacterial LRTI  N = 58 | p-value* |
| --- | --- | --- | --- |
| G-CSF | 218.3 (37.1 – 787.1) | 56.2 (15.3 – 272.2) | 0.268 |
| IL-1β | 213.7 (12.9 – 500.0) | 74.8 (11.2 – 1088.5) | 0.360 |
| IL-6 | 49.4 (7.9 – 201.3) | 34.4 (2.8 – 128.4) | 0.401 |
| IL-8 | 3142.3 (1332.0 – 12399.1) | 2348.6 (348.3 – 6190.0) | 0.393 |
| IP-10 | 34.6 (11.6 – 1632.6) | 18.5 (6.3 – 221.4) | 0.456 |
| MCP | 19.9 (5.9 – 196.2) | 10.6 (2.1 – 94.9) | 0.268 |
| MIP-1α | 23.2 (3.3 – 79.1) | 6.3 (1.3 – 35.5) | 0.352 |
| TRAIL | 147.5 (75.5 – 234.3) | 53.3 (11.8 – 319.5) | 0.336 |
| ^*Mann-Whitney U test with Benjamani and Hochberg correction for multiple comparisons^ | | | |

**Table S9. Receiver-operating characteristic (ROC) analysis of plasma IL-6 and CRP for discriminating bacterial versus non-bacterial LRTI (RASCALS cohort).**
Area under the curve (AUC), optimal cut-off values, sensitivity, specificity, positive predictive value (PPV), and negative predictive value (NPV) are reported. CRP achieved the highest diagnostic accuracy (AUC 0·81), with IL-6 also showing discriminatory value (AUC 0·74).

**List 1.** The intersection of differential expressed genes in DESeq2 and edgeR.

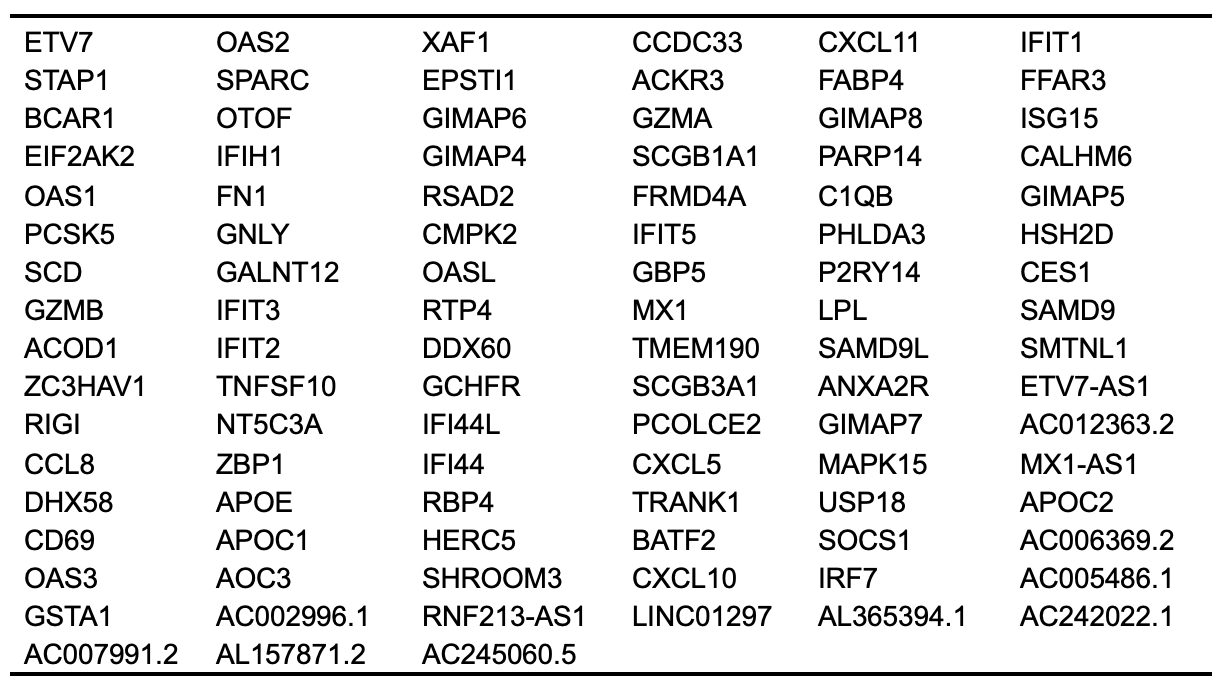

**List 2.** Key proteins from the LRTI network in each centrality measure.

Eigenvector centrality:

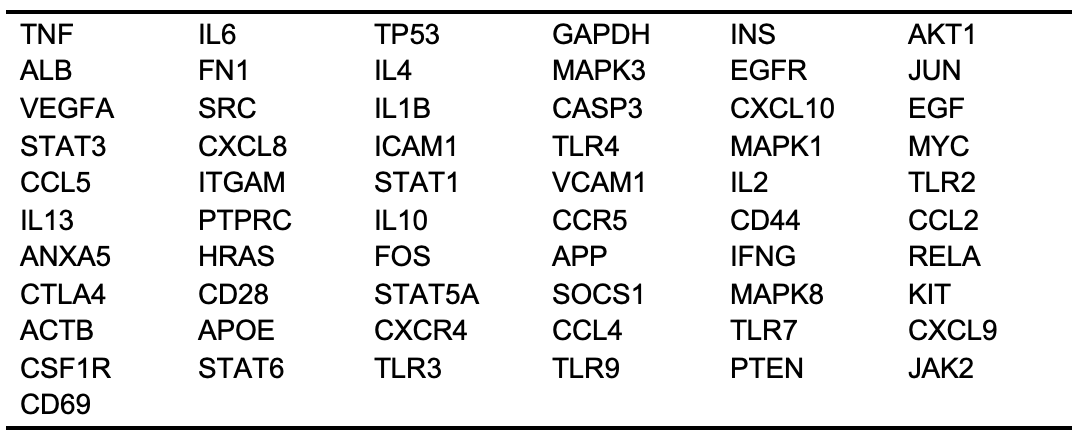

Degree centrality:

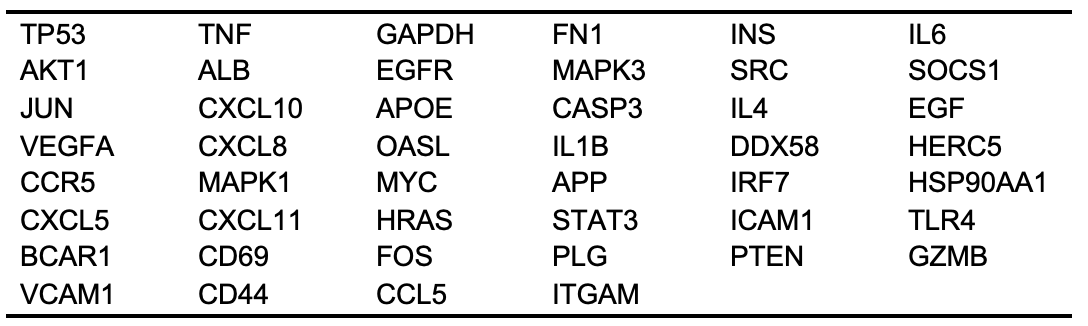

Betweenness centrality:

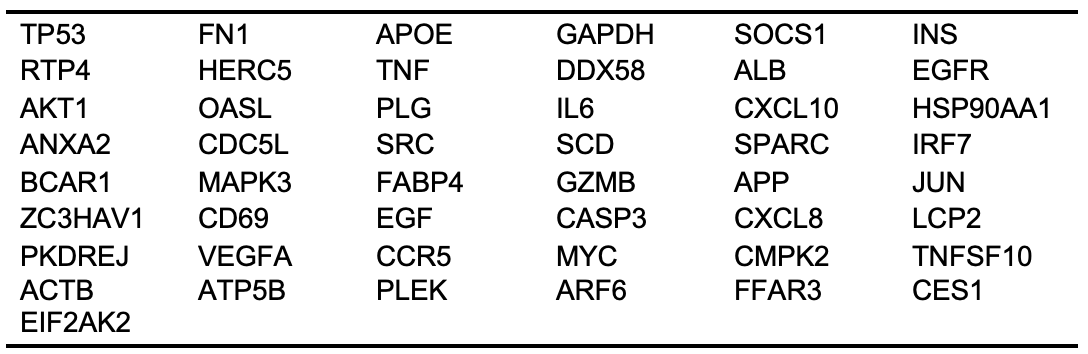

RWR:

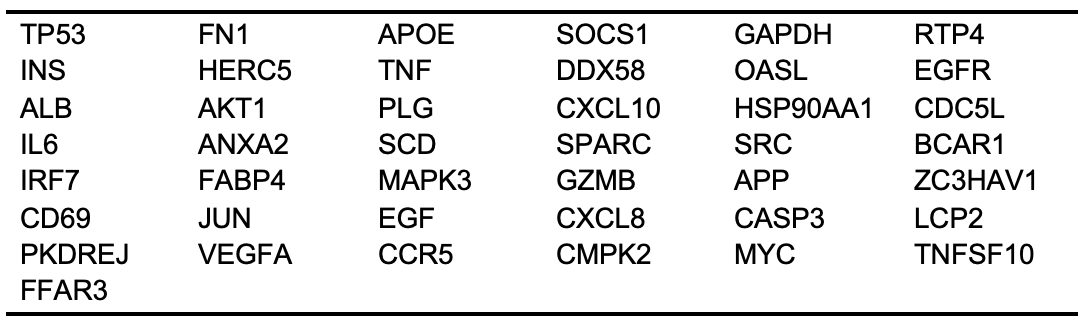

**List 3.** Key proteins from the RSV-host network in each centrality measure.

Eigenvector centrality:

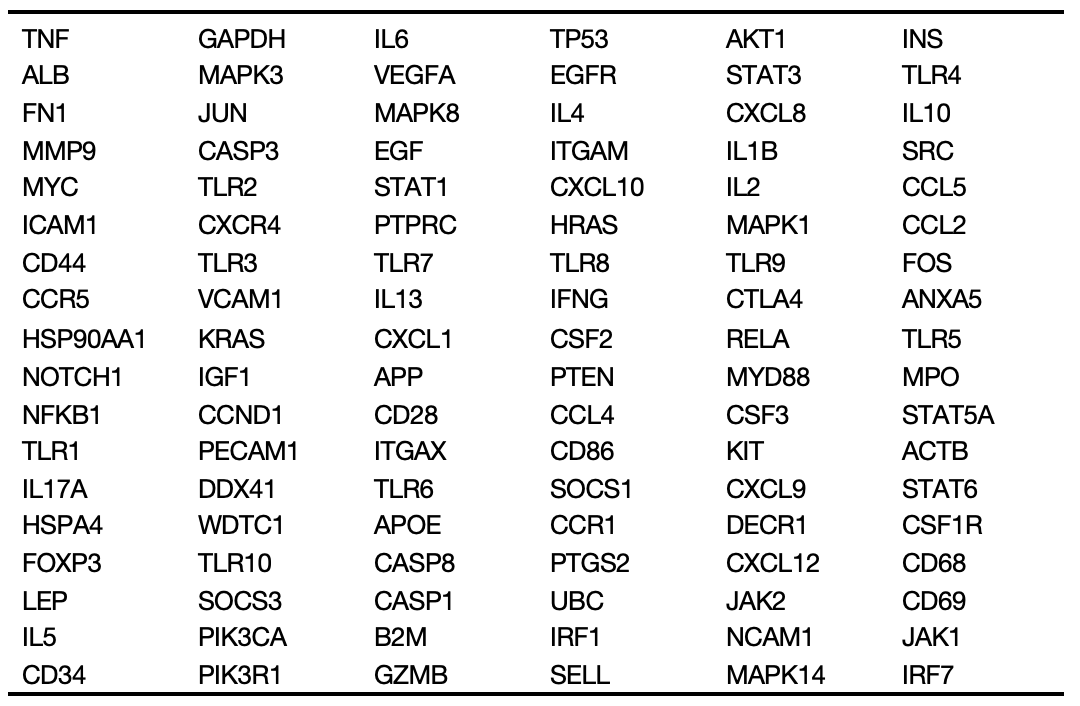

Degree centrality:

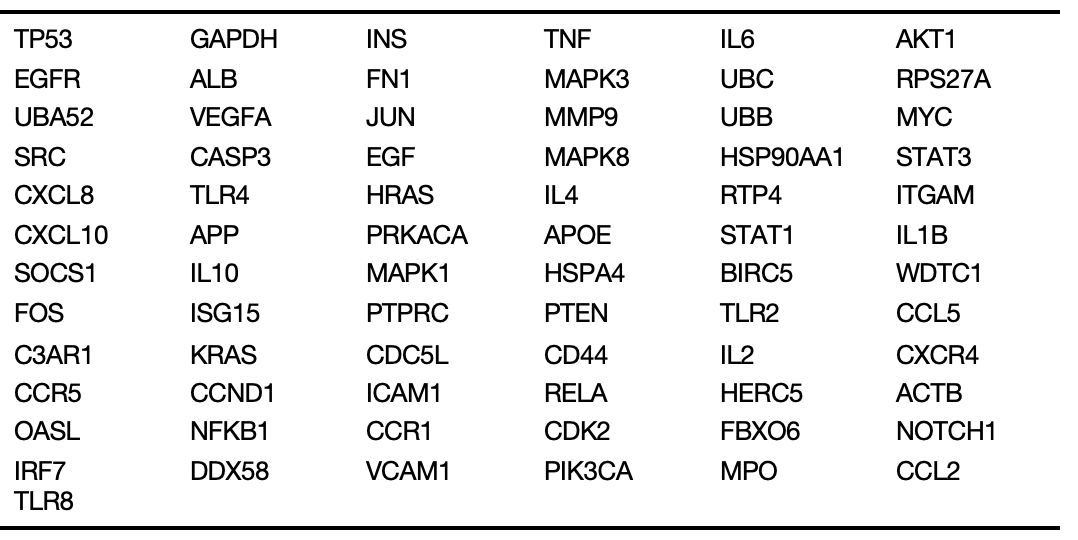

Betweenness centrality:

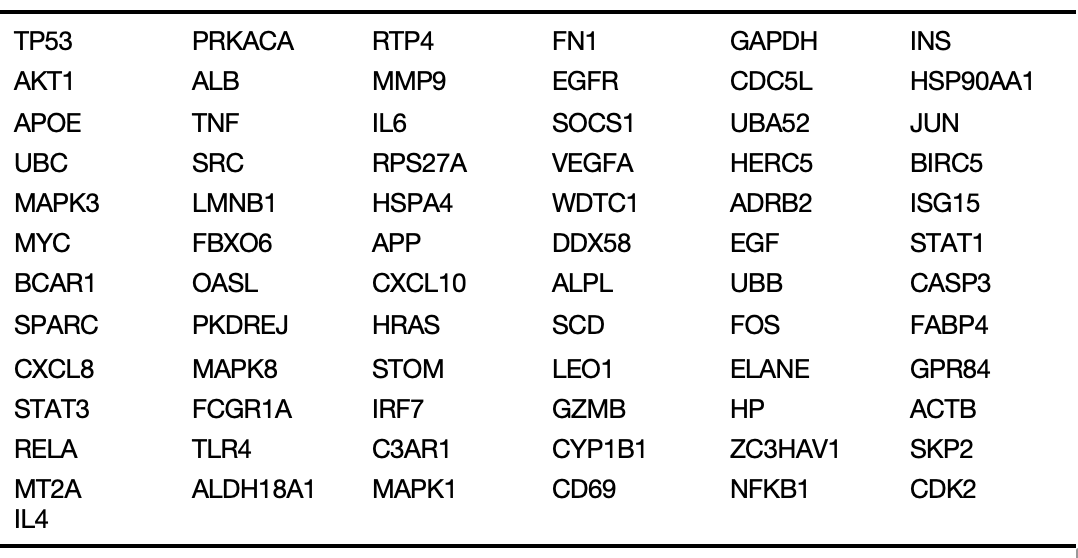

RWR:

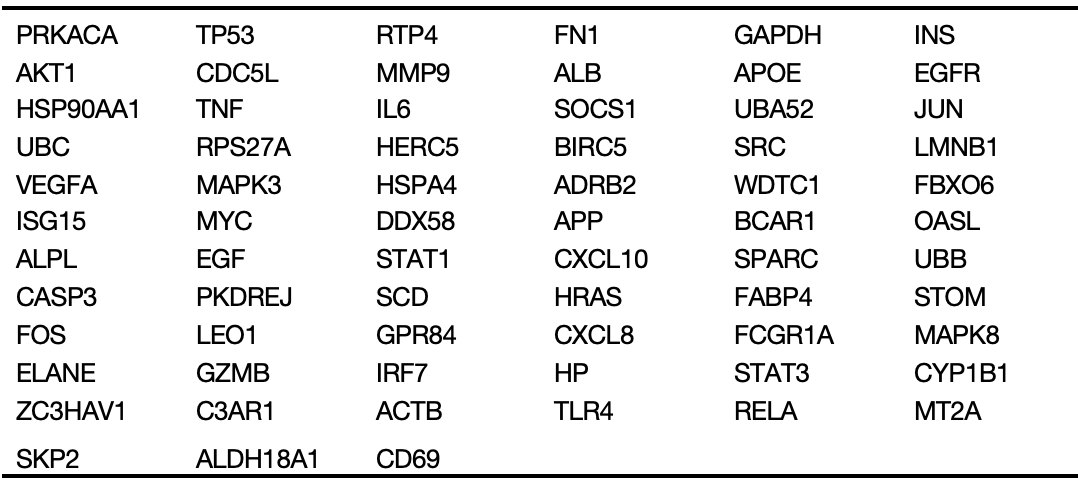
